# Improving estimation efficiencies for family-based GWAS by integrating large external data

**DOI:** 10.64898/2025.12.26.25343073

**Authors:** Zixuan Wu, Yunqi Yang, Aabesh Bhattacharyya, Matthew Stephens, Jingshu Wang

**Affiliations:** Department of Statistics, University of Chicago, Chicago, IL 60637, USA; Committee on Genetics, Genomics and System Biology, University of Chicago, Chicago, IL 60637, USA; Department of Human Genetics, University of Chicago, Chicago, IL 60637, USA

**Keywords:** within-family GWAS, summary statistics, calibration, direct genetic effects, genetic nurture, Mendelian randomization, variance reduction, assortative mating

## Abstract

Family-based genome-wide association studies (GWAS) can separate direct from indirect genetic effects such as genetic nurture, population stratification, and assortative mating, yet these designs often suffer from limited statistical power because large samples of genotyped trios and sibships are rare. We introduce a calibration framework that improves the efficiency of within-family GWAS by integrating three readily available summary statistics for each SNP: the within-family association, the corresponding population-based estimate from the same family sample, and an external population-based estimate from a large GWAS. The method does not require individual-level data and is compatible with generalized linear models used for both continuous and binary traits. Theoretical results show that calibration can reduce variance by up to fifty percent in trio designs and up to twenty-five percent in sibling designs, equivalent to doubling the effective sample size for trios. Simulations confirm the accuracy and unbiasedness of the calibrated estimator, and applications to UK Biobank family data demonstrate substantial precision gains and improved downstream Mendelian Randomization inference. Analysis of published within-sibship GWAS summary statistics further illustrates that the approach can be applied directly to publicly available data. Together, these results show that calibration provides a practical and powerful way to enhance family-based genetic analyses.

## 1 Introduction

Genome-wide association studies (GWAS) have identified thousands of genetic variants linked to complex human traits [Buniello et al., 2019; Visscher et al., 2017]. Traditional GWAS, conducted in large cohorts of unrelated individuals, estimate marginal SNP–trait associations that may reflect not only direct genetic effects but also indirect effects, such as genetic nurturing where parental genotypes influence offspring outcomes through environmental pathways [Kong et al., 2018]. These associations may also be confounded by linkage disequilibrium (LD) and demographic factors, including population stratification and assortative mating [Howe et al., 2022; Veller and Coop, 2024].

Separating direct from indirect genetic effects is crucial for identifying causal variants and understanding the environmental pathways through which genetic variation influences phenotypes [Balbona et al., 2021; Evans et al., 2019; Hegemann et al., 2025; Tubbs et al., 2020]. Within-family GWAS provides a principled solution by leveraging random Mendelian segregation. Conditional on parental genotypes, transmitted alleles are completely randomized, and differences between siblings are also independent of shared familial confounding [Howe et al., 2022; Young et al., 2019]. As a result, within-family GWAS provides estimates of direct genetic effects that are robust to indirect genetic effects, population stratification, and assortative mating. Such bias-robust estimates are also valuable for downstream analyses such as within-family Mendelian randomization (MR), which uses within-family associations for both exposures and outcomes to mitigate violations of the core assumption that genetic instruments are independent of environmental confounders [Brumpton et al., 2020; Davies et al., 2019].

Despite these advantages, within-family analyses are constrained by the practical difficulty of collecting large numbers of genotyped families. While biobank-scale studies include hundreds of thousands of participants, the subset with suitable family structures is much smaller. For example, the UK Biobank includes nearly 500,000 genotyped participants yet only 1,066 complete trios and 22,666 sibling pairs [Bycroft et al., 2018], leading to substantially larger standard errors than conventional population-based GWAS.

To address this efficiency gap, we propose a calibration framework that combines noisy but unbiased within-family GWAS estimates with more precise, though potentially biased, estimates from large-scale population-based GWAS. Our method uses three summary statistics: the within-family SNP–trait association, the corresponding population-based association from the same family sample, and the population-based association from large-scale GWAS data. Unlike recent approaches that improve efficiency by imputing parental genotypes [Guan et al., 2025; Young et al., 2022], our method does not require individual-level data or imputation. It operates directly on summary statistics and involves modest computational overhead, making it both practical and scalable. This framework enhances the efficiency of within-family analyses and facilitates more powerful applications, such as within-family MR.

## 2 Materials and methods

### 2.1 Overview of the method

Our method improves the efficiency of within-family GWAS estimates by combining two data sources: an internal cohort with family genotype data, which yields unbiased but noisy estimates of direct genetic effects, and an external large-scale population GWAS, which provides precise but potentially biased estimates.

Our workflow has three steps (Figure 1). First, for each SNP *j*, we obtain three association estimates: (i) 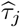, the within-family SNP–trait association estimated from the internal data adjusting for covariates ***C*** and family genotype ***F*** _*j*_; (ii) 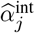, the population-based marginal association from the same family cohort adjusting only for ***C***; and (iii) 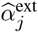, the population-based marginal association from a large external GWAS. We then assess and correct for scale differences between 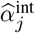 and 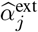 to ensure their comparability, and form the contrast 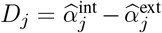. Finally, we construct a calibrated within-family estimate through 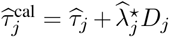, where the weight 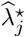 is chosen to minimize variance of 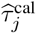. This construction follows the general calibration principle used in survey sampling [Cochran, 1977] and the control variates idea in Monte Carlo estimation [Owen, 2013]: an unbiased but noisy estimator can be improved by adding a mean-zero auxiliary term. In our setting, the difference *D*_*j*_ serves as that auxiliary quantity. The remainder of this section details the models, calibration, and inference procedures, and provides theoretical results on efficiency gains.

**Figure 1:**
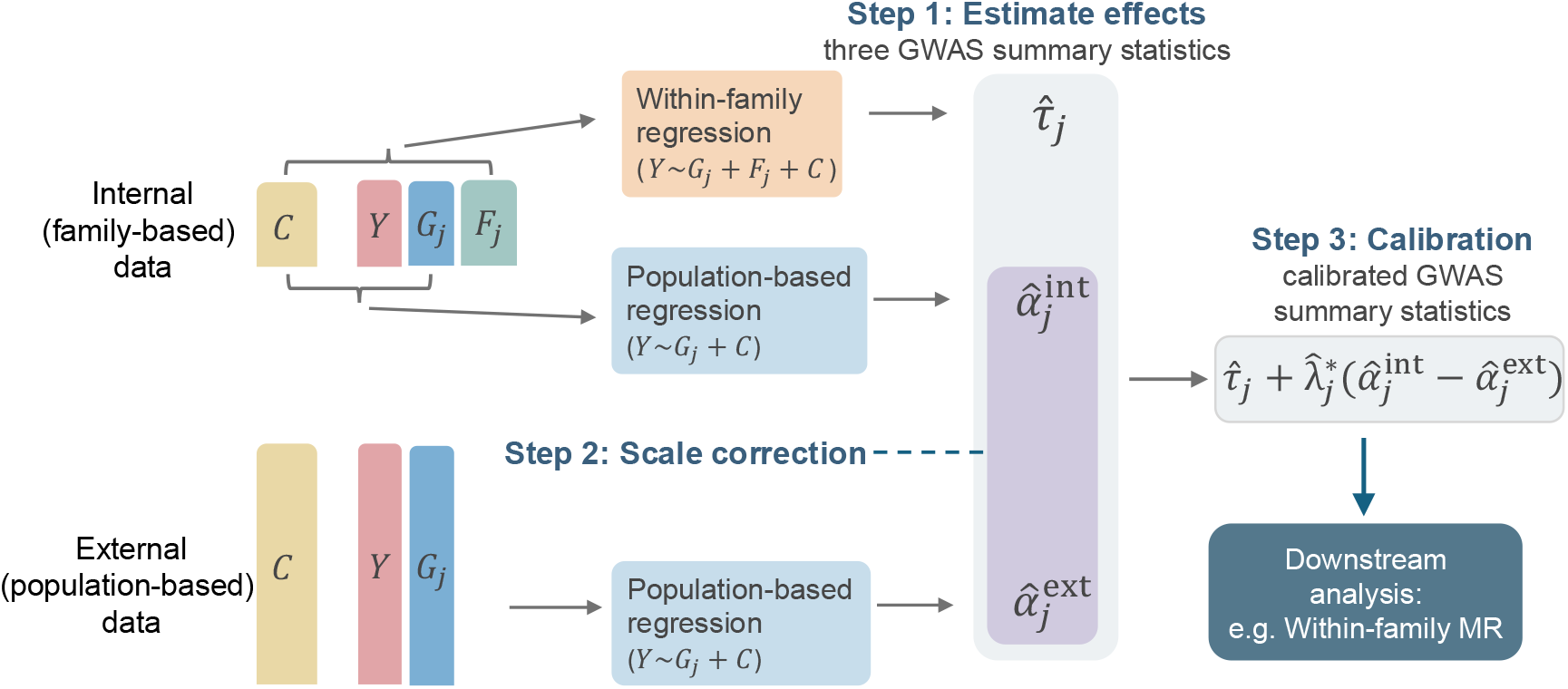
Workflow of the calibration method. Internal data provides within-family and population-based estimates, and external GWAS provides large-scale population-based estimates. These are aligned and combined to produce a calibrated within-family estimator.

### 2.2 Regression models

We now formalize the notation introduced in the overview. Our approach is derived from a single-locus model, where SNP effects are estimated one at a time. Consider an individual *i* and SNP *j*. Let *Y*_*i*_ denote the phenotype of individual *i, G*_*ij*_ the genotype at SNP *j*, and ***C***_*i*_ a vector of observed covariates including the intercept and other commonly adjusted factors such as age, sex, and population principal components. Let ***F*** _*ij*_ denote family genotype information for SNP *j* used to control shared familial effects, for example, parental genotypes or the sum of sibling genotypes. Our target is the direct genetic effect *τ*_*j*_ of SNP *j* on the phenotype.

In the internal family cohort, we estimate *τ*_*j*_ by fitting a generalized linear model that conditions on family genotype:

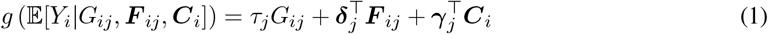

where *g*(·) is the link function. Our framework accommodates general link functions, so the derivations below are not tied to a specific choice. Common choices are the identity link for continuous traits and the logit link for binary traits.

Table 1 summarizes common choices for the family genotype ***F*** _*ij*_ across study designs. For example, with both parents genotyped, ***F*** _*ij*_ can be the sum of maternal and paternal genotypes [Young et al., 2022] (equivalent to adjusting for the non-transmitted alleles *NT*_*ij*_, in which case *τ*_*j*_ is the difference between the coefficients of *G*_*ij*_ and *NT*_*ij*_ [Kong et al., 2018]), or the vector of the two parental genotypes [Brumpton et al., 2020; Wu et al., 2021]. In sibling-based designs, we take 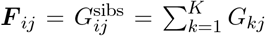, the sum of sibling genotypes within the family [Howe et al., 2022]. When *K* = 2 and a linear regression model is used, fitting (1) with 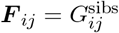 in equivalent to the two-sibling difference regression, where the difference of the outcome between two siblings *Y*_1_ − *Y*_2_ is regressed on the difference of the genotype between two siblings *G*_1*j*_ − *G*_2*j*_ [Brumpton et al., 2020]. Further details are provided in the Supplementary Text.

**Table 1:** Common choices for family genotype information and the implied direct effect estimand. coef(·) denotes the regression coefficient on the indicated predictor. For parent–offspring designs, conditioning on the non-transmitted allele (*NT*_*ij*_) is algebraically equivalent to conditioning on the sum of parental genotypes. For sibling designs, using the sum of sibling genotypes is equivalent to the two-sibling difference formulation when *K* = 2. Thus, all cases are covered in model (1).

| Study design | Family genotype $\mathbf{F}_{ij}$ | Direct effect $\tau_j$ |
| --- | --- | --- |
| Parents + offspring | $G_{ij}^{\text{pa}} = G_{ij}^m + G_{ij}^f$ | $\text{coef}(G_{ij})$ |
| Parents + offspring | $NT_{ij}$ | $\text{coef}(G_{ij}) - \text{coef}(NT_{ij})$ |
| Parents + offspring | $(G_{ij}^m, G_{ij}^f)$ | $\text{coef}(G_{ij})$ |
| Mother + offspring | $G_{ij}^m$ | $\text{coef}(G_{ij})$ |
| Siblings ($K \geq 2$) | $G_j^{\text{sibs}} = \sum_{k=1}^K G_{kj}$ | $\text{coef}(G_{ij})$ |
| Siblings (two-sibling difference) | Regress $(Y_1 - Y_2)$ on $(G_{1j} - G_{2j})$ | $\text{coef}(G_{1j} - G_{2j})$ |

For comparison, we also consider the standard population-based GWAS model,

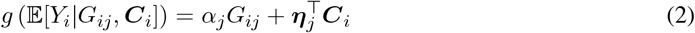

which regresses the outcome on the SNP and common covariates without family adjustment. Fitting model (2) on the internal data yields the internal population-based estimate 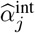, while fitting the same model on a large external GWAS provides a more precise estimate 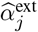 for *α*_*j*_.

We assume the family-adjusted model in Eq. (1) is correctly specified so that *τ*_*j*_ identifies the direct genetic effect. This relies on random Mendelian segregation, adequate adjustment for indirect familial effects via ***F*** _*ij*_, and a correct link *g*(·). Additional assumptions for calibration and summary-statistics inference are stated in later sections.

### 2.3 Scale correction and calibration principle

The calibration estimator [Chen and Chen, 2000; Yang and Ding, 2019] takes the form

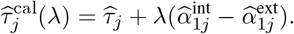

In principle, this estimator remains unbiased for *τ*_*j*_ for any fixed *λ*, since both 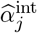 and 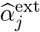 estimate the same target *α*_*j*_, making their difference mean zero. This requires that model (1) is correctly specified in both datasets and that genotype distributions are comparable across the internal and external cohorts (see Supplemental Text for details).

In practice, however, phenotype scaling and analytic pipelines often differ across cohorts, leading to systematic scale discrepancies between 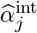 and 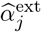. To correct for this, we introduce a multiplicative scale factor *s* and assume 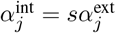, where 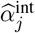 and 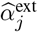 are unbiased estimators of their respective targets. We estimate *s* by regressing 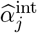 on 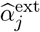 across LD-independent SNPs, using an MR method such as GRAPPLE [Wang et al., 2021], and take the slope estimate as 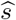. The external estimates are then rescaled to 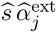 (redefined as the new 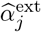) prior to calibration. With this scale alignment, the contrast 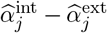 is centered at zero, ensuring unbiasedness of the calibrated estimator and enabling variance reduction through the choice of *λ*.

### 2.4 Theoretical variance reduction

Under weak local dependence across individuals, a joint central limit theorem holds for 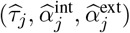. Minimizing the variance of 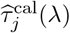 gives the optimal weight

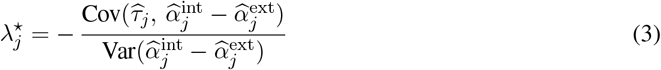

and the corresponding variance

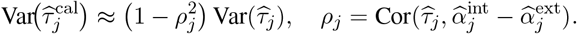

Thus, the proportion of variance reduction relative to the uncalibrated estimator is 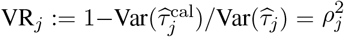.

To obtain closed-form approximations of VR_*j*_, we assume that per-SNP direct and indirect effects are small relative to trait variability, that individuals in the external data are independent, and that families are independent in the internal data (for formal mathematical statements of the theoretical results, see the Supplementary Text). We denote by *N*_int_, *N*_ext_ and *N*_share_ the sizes of the internal, external, and overlapping samples between the internal and external data.

For parent–offspring designs with both parents genotyped, the variance reduction is approximately

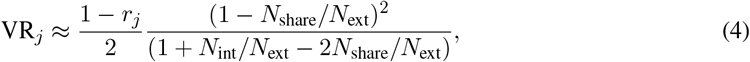

where *r*_*j*_ = Cor(*G*_*ij*_, *NT*_*ij*_) is the correlation between transmitted and non-transmitted alleles. When *r*_*j*_ = 0 (random mating), and the external cohort is much larger than the internal, the variance reduction is about 50%, corresponding to effectively doubling the internal sample size.

In sibling-based designs with the sibling size *K* = 2, the variance reduction is approximately

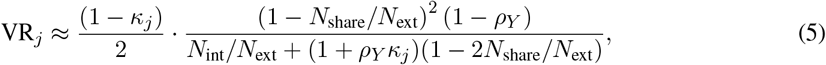

where 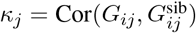 and 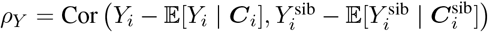 is the phenotypic corre-lation between two siblings after adjusting for covariates. When *κ*_*i*_ ≈ 0.5 (random mating) and with large external data, this simplifies to (1 − *ρ*_*Y*_)*/*(4 + 2*ρ*_*Y*_), which decreases from roughly 25% when siblings are uncorrelated (*ρ*_*Y*_ = 0) to 0 as phenotypic correlation approaches one.

### 2.5 Calibration with summary statistics

We next show how to estimate the optimal weight 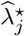 only using the summary statistics instead of individual-level data. We rely on the three association estimates introduced above 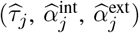, along with their reported standard errors. As in Section 2.4, we assume that per-SNP direct and indirect effects are small relative to trait variability.

To make calibration feasible with only these quantities, we further assume that the correlation structure among family members is stable across SNPs. Formally, for ***X***_*ij*_ = (*G*_*ij*_, ***F*** _*ij*_) combining the genotype and family information for individual *i* at SNP *j*, we require that

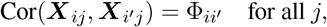

where 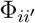 depends on the relationship between *i* and *i*^*′*^ (e.g., siblings, parent–child, unrelated) but not on the SNP itself. Intuitively, this means that relatedness patterns arising from assortative mating and family structure are consistent across the genome. Under this assumption, the correlations 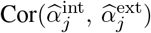 and 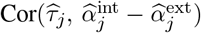 approximately constant across all SNPs. (see Supplementary Text for derivations).

In practice, to estimate the shared 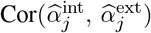, we compute the median of the standardized products of the internal and external *z*-scores across all SNPs. Unlike Wang et al. [2021] which estimates this correlation using the mean over empirical-null SNPs, we use a median-based estimator over all SNPs for greater robustness and computational simplicity.

To estimate the shared 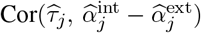, we use the formula

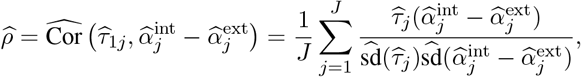

where *J* is the number of SNPs. This leads to a practical form of the calibrated estimator:

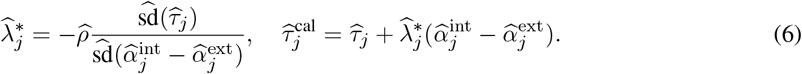

By construction, the calibrated estimator is approximately normal: 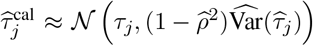 so a (1 − *α*) Wald interval is 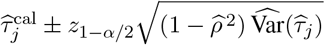. Relative to the uncalibrated interval, this shrinks in length by a factor of 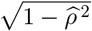.

Although the stable correlation assumption is convenient, calibration can also be carried out allowing heterogeneous correlation across SNPs for sibling and trio data, at the cost of stronger modeling assumptions and more involved calculations. These extensions are described in the Supplementary Text and expand the usability of our calibration approach.

#### Individual-level versus summary-level calibration

When individual-level genotype and phenotype data are available, the optimal weight 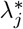 in Eq. (3) can be estimated directly from regression, together with the variance of this contrast. We refer to this as individual-level calibration.

The summary-level calibration estimates these quantities only from the three summary statistics as described above. Both approaches have the same form, the difference lies only in how 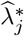 is obtained.

### 2.6 Simulation Setup

We conducted simulation studies to evaluate the efficiency and reliability of the calibration procedure under settings that mimic commonly used trio- and sibling-based designs.

#### Genotype generation

We simulated 100 independent biallelic SNPs. For each SNP, the allele frequencies were sampled from the empirical distribution in the UK Biobank (Neale Lab, restricted to the interval [0.01, 0.99]). Parental genotypes 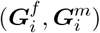 were generated under Hardy–Weinberg equilibrium, and offspring genotypes ***G***_*i*_ were created through random Mendelian transmission, i.e., by inheriting one allele from each parent at every locus independently. This process produced either a trio structure (mother, father, one child) or a sibling structure (mother, father, two children).

#### Phenotype generation

To induce indirect genetic effects, we first simulated parental phenotypes using an additive model 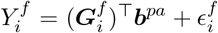 and 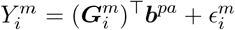, where the parental effect sizes ***b***^*pa*^ were drawn from multivariate normal distribution *N* (0, *I*), and the errors 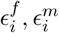 where draw from *N* (0, *σ*^2^). These parental traits contribute to indirect genetic effects on the offspring phenotypes.

Offspring phenotypes incorporated both direct and indirect effects, and were generated as

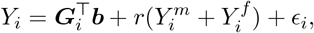

for continuous traits, where ***b*** ∼ *N* (0, *I*), *ϵ*_*i*_ ∼ *N* (0, *σ*^2^), and *r* controls the strengh of indirect parental effects. Binary traits were generated using a liability-threshold transformation of the continous traits (see more details in the Supplementary Text).

#### Family designs and sample sizes

Trio families were generated by simulating one offspring from each parent pair and sibling families were generated by simulating two offspring from each parental pair. Each simulation included an internal family-based cohort with 10,000 independent families and an external population-based cohort with 100,000 unrelated individuals. The effect size vectors ***b***^*pa*^ and ***b*** were generated once and kept fixed across all simulation replicates.

#### Simulation configurations

We varied key aspects of the data-generating process, including (i) the strength of indirect effects *r*, (ii) the residual variance *σ*^2^ (which determines the phenotypic correlation between siblings), and (iii) the family structure (trios vs. siblings). For each configuration, we generated 500 replicates and computed within-family, internal population-based, and external population-based association estimates before applying the calibration procedure.

### 2.7 Real data analyses

#### 2.7.1 UK Biobank family-based analysis

We applied our calibration approach to family-based genotype data from the UK Biobank, a population-based cohort with extensive genotype and phenotype information for more than 500,000 participants aged 40–69 at recruitment [Bycroft et al., 2018; Sudlow et al., 2015]. We analyzed six traits: body mass index (BMI; field 21001), diastolic blood pressure (DBP; 4079), systolic blood pressure (SBP; 4080), diabetes (2443), current tobacco smoking (1239) and years of education (derived from field 6138 following Howe et al. [2023]). For downstream MR analyses, BMI was treated as the exposure and the other traits as outcomes.

##### Family identification

Family structures were inferred using UK Biobank kinship data. Parent–offspring and full-sibling pairs were first identified using a kinship coefficient between 2^−5*/*2^ and 2^−3*/*2^. Then, parent–offspring pairs were defined by IBD0 ≤ 0.0012, whereas full siblings were defined by IBD0 between 0.0012 and 0.365 [Bycroft et al., 2018; Manichaikul et al., 2010]. This yielded 6,263 parent–offspring pairs, 22,646 sibling pairs, and 1,172 complete trios. Non-overlapping families were further reconstructed using a breadth-first search on the kinship graph, producing 4,958 parent–offspring families and 20,128 sibling families. Individuals without close relatives (data field 22021 = 0) were used as the external sample.

##### Generation of within-family and population-based GWAS

For each trait, we generated three sets of association statistics from the UK Biobank: (i) within-family estimates, (ii) internal population-based estimates, and (iii) external population-based estimates. The within-family and internal population-based GWAS were computed using only individuals belonging to reconstructed family units (trios or sibling families), whereas the external population-based GWAS was generated from the unrelated individuals in UK Biobank.

Genotypes were extracted from the UK Biobank BGEN files using PLINK2 after applying standard variant-level QC: minor allele frequency ≥ 1%, Hardy–Weinberg Equilibrium p-value ≥ 10^−6^, biallelic SNPs only and removal of duplicated variants. All traits were standardized, and all three association statistics were estimated using linear regression adjusting for standardized age, age_2_, sex, and the first 10 principal components.

##### MR setup

To construct genetic instruments for BMI, we performed LD clumping using the 1000 Genomes European reference panel [Consortium et al., 2015] and selected SNPs associated with BMI with p-values less than 10^−8^ in the GIANT consortium GWAS [Locke et al., 2015; Wood et al., 2014]. This yielded 57 approximately independent variants. Causal effects were estimated using MR.RAPS [Zhao et al., 2019, 2020] applied to the within-family estimates, internal population-based estimates, and their calibrated versions.

#### 2.7.2 Calibration using published within-sibship GWAS summary statistics

We further evaluated our method using the within-sibship GWAS summary statistics reported by Howe et al. [2022], which analyzed 178,076 siblings from 77,832 sibships across 19 studies. These data contain both population-based and within-sibship association estimates only using these siblings across numerous traits and were used in the original study for within-family MR of BMI, height, and several cardiometabolic outcomes. Because family-based GWAS are typically underpowered, MR estimates derived from these data often have wide confidence intervals. Our goal was to assess whether calibration can improve efficiency when applied directly to published summary statistics.

##### Calibration

For each phenotype, we applied our calibration procedure to the full set of available SNPs. Additionally large-scale population-based GWAS summary statistics are served as external references. Scale correction between the internal and external population-based GWAS estimates was performed using GRAPPLE [Wang et al., 2021], following Section 2.3.

##### MR setup

SNP selection and LD clumping followed the same procedure used in our UK Biobank analysis (Section 2.7.1), using separate trait-specific SNP-selection files and p-value cutoff of 5 × 10^−8^. We applied both inverse-variance weighting (IVW, Burgess et al. [2013]), to enable comparison with the original Howe et al. analysis, and MR.RAPS, which provides more stable estimates when instruments are weak or noisy.

Complete details of the specific datasets used as external population-based GWAS and SNP-selection files are provided in the Supplementary Materials.

## 3 Results

### 3.1 Simulation results

We first evaluated the performance of the calibrated estimator simulations of trio and sibling designs for both continuous and binary traits. Across four settings, trio continuous, trio binary, sibling continuous, and sibling binary, the estimated variance of the calibrated estimator closely matched its empirical variance across 500 simulated replicates (Figure 2a). Linear regression was used for continuous traits and logistic regression for binary traits. Points fell tightly along the diagonal, demonstrating that the variance formula underlying our calibrated summary statistics is accurate.

**Figure 2:**
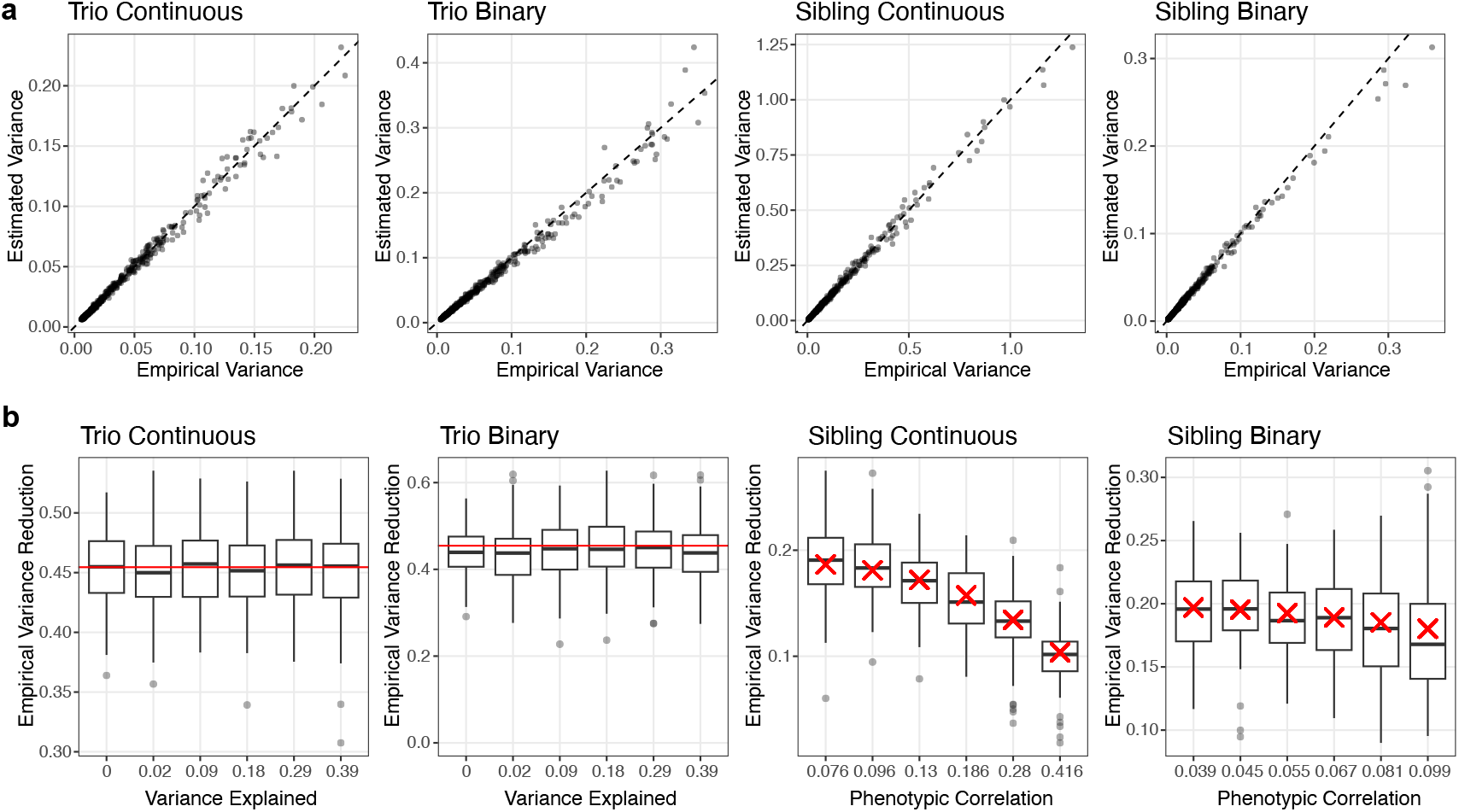
Simulation results. a) Comparison of the estimated variance of the calibrated estimator with its empirical variance across 500 simulation replicates. Each black dot represents one SNP, and the diagonal line denotes *y* = *x*. b) Empirical variance reduction of the calibrated estimator relative to the uncalibrated estimator for 100 SNPs under linear (continuous-trait) and logistic (binary-trait) regression in trio and sibling designs. Each black dot represents one SNP; boxplots summarize the distribution (median, interquartile range, and whiskers showing the central 1.5 IQR range) across SNPs. Red lines or crosses denote the corresponding theoretical variance reduction.

We next examined variance reduction for each SNP by comparing the empirical variance of the calibrated and uncalibrated estimators (Figure 2b). In the trio designs, the strength of indirect parental effects is quantified on the x-axis using the ratio 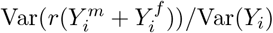, and and the empirical reductions were tightly centered around a constant theoretical value following Equation (4) regardless of the strength of the indirect effects (approximately 45% for an internal-to-external sample-size ratio of 0.1). In the sibling designs, variance reduction decreased with increasing phenotypic correlation between siblings, as predicted in Equation (5), averaging roughly 20% when the correlation was near zero and 10% when it approached 0.4.

Finally, we repeated these analyses using only GWAS summary statistics to construct the calibrated estimator, and observed nearly identical results (Supplementary Figure S1). This demonstrates that summary-level calibration performs equivalently to individual-level calibration, further supporting the practical utility of our approach in settings where only summary statistics are available.

### 3.2 Calibration for UK Biobank Family-Based GWAS

Within-family designs help separate direct genetic effects from residual familial influences, making accurate estimation particularly important for complex traits such as blood pressure, diabetes, education, and smoking. We applied our calibration framework to the UK Biobank family-based data to evaluate its performance in real-world settings. For each of the six traits, we generated within-family and population-based association estimates and then applied both the individual-level and summary-level calibration procedures described in Section 2.5.

Figure 3a–b summarizes variance reduction across 1,000 randomly sampled SNPs. In trio data, calibration reduced the variance of family-based estimates by roughly 50% for all traits, matching the theoretical expectation in Equation (4). In sibling data, variance reduction also aligned with theory and varied by trait, reflecting differences in phenotypic correlation between siblings: traits with weaker within-family correlation (for example, diabetes or current smoking) exhibited larger variance reductions (≈ 20%), whereas highly correlated traits such as education showed more modest gains (Equation (5) showing that VR_*j*_ decreases monotonely with *ρ*_*Y*_). Calibration performed using summary statistics produced nearly identical variance reduction as calibration performed with individual-level data, consistent with our simulation findings.

**Figure 3:**
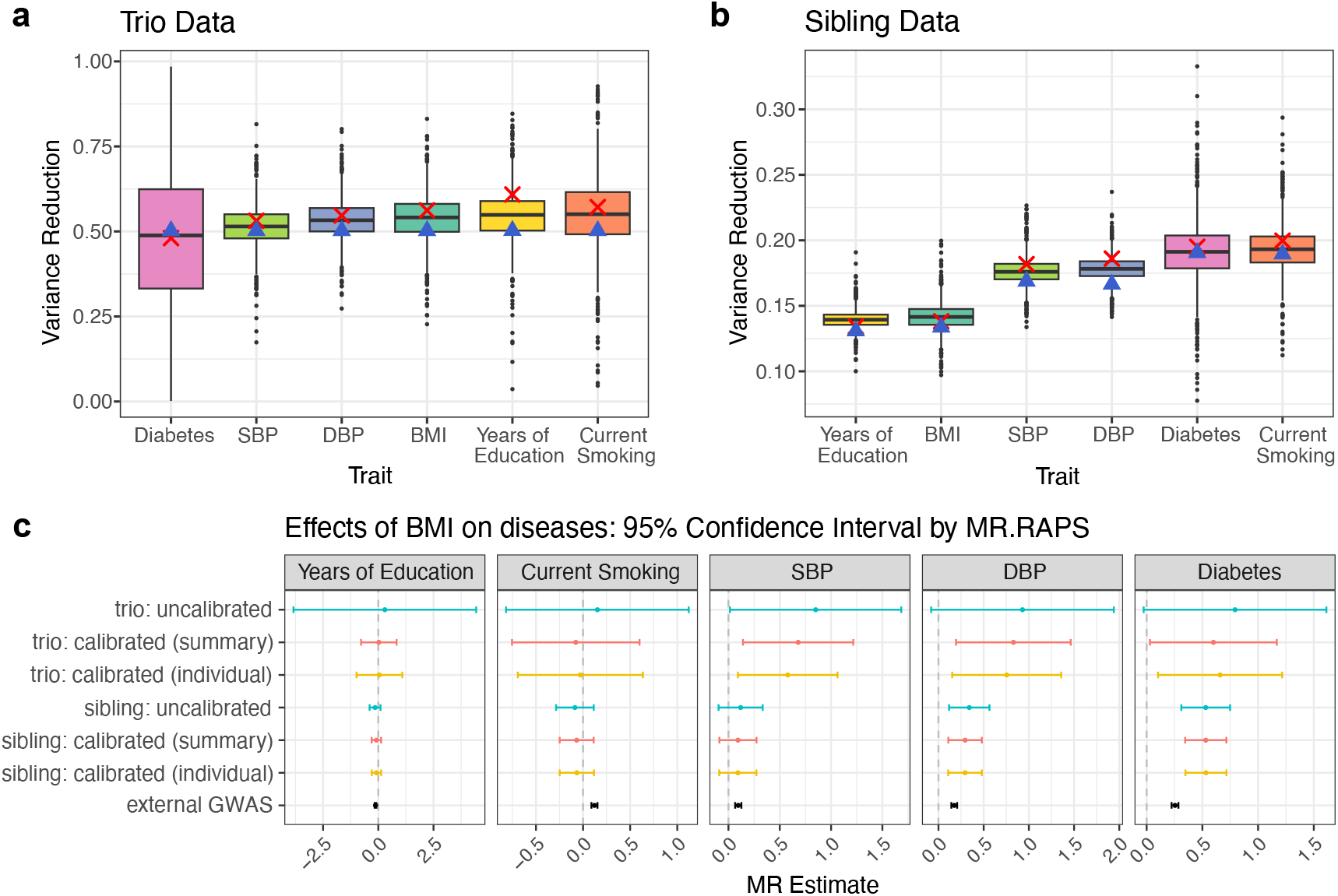
Calibration applied to UK Biobank traits. Estimated variance reduction when used on a) trio and b)sibling data. The boxplots are based on 1,000 randomly sampled SNPs and summarize the distribution of estimated variance reduction under the trio and sibling designs (median, interquartile range, and whiskers showing the central 1.5 IQR range). Red crosses indicate the variance reduction obtained from the summary-level calibration (identical across all SNPs), and blue triangles denote the corresponding theoretical values. c) Mendelian randomization results using MR.RAPS on UK Biobank data. Error bars represent 1.96 times the standard error returned by the corresponding MR method.

Additional diagnostic results further support both the accuracy and unbiasedness of the calibrated estimator. First, for each trait, the distribution of the difference between calibrated and uncalibrated effect estimates was tightly centered at zero across all SNPs (Figure S2a), with particularly concentrated distributions in sibling data where residual noise is smaller. Second, the estimated calibration weights 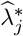 obtained from summary-statistics calibration aligned extremely well with those obtained from individual-level calibration (Figure S2b), indicating that summary-only calibration yields similar adjustment as individual-level implementation. Together, these results confirm that our method behaves as theoretically predicted and that summary-level calibration is accurate and practically reliable.

Finally, we evaluated downstream MR using BMI as the exposure (Figure 3c). MR using external population-based GWAS suggested significant associations between BMI and all five outcomes. When we replaced the population-based associations with within-family estimates, the MR estimates for education and current smoking were smaller and became insignificant, consistent with prior work indicating that these traits may be influenced by indirect genetic effects such as genetic nurture [Davies et al., 2019; Howe et al., 2022; Kong et al., 2018]. Calibration substantially improved precision in the trio analyses: uncalibrated within-family MR lacked power to detect the causal effects of BMI on diabetes, SBP, and DBP, whereas calibrated trio MR recovered significant and directionally consistent estimates. Sibling-based MR exhibited similar patterns, with smaller efficiency gains. Importantly, MR based on summary-calibrated estimates closely matched MR based on individual-level calibration, supporting that summary-level calibration is robust for downstream causal inference.

### 3.3 Calibration for Published Within-Sibship GWAS Summary Statistics

We next applied our method to the within-sibship GWAS summary statistics of Howe et al. [2022], which include 178,076 siblings from 77,832 sibships across 19 cohorts. This analysis illustrates that our calibration framework can be applied directly to publicly available summary data, without requiring individual-level genotypes or phenotypes.

Figure 4a summarizes variance reduction across all traits. Calibrated within-family effect estimates showed 10–20% variance reduction, with the magnitude reflecting trait-specific sibling phenotypic correlations: traits with weaker within-family correlation (for example, SBP or age at first birth) benefited most, whereas traits with stronger correlation (for example, years of schooling or height) showed more modest gains. Although the absolute reduction is smaller than in the UK Biobank trios, it is essentially “free”, requiring only external population GWAS and minimal computation.

**Figure 4:**
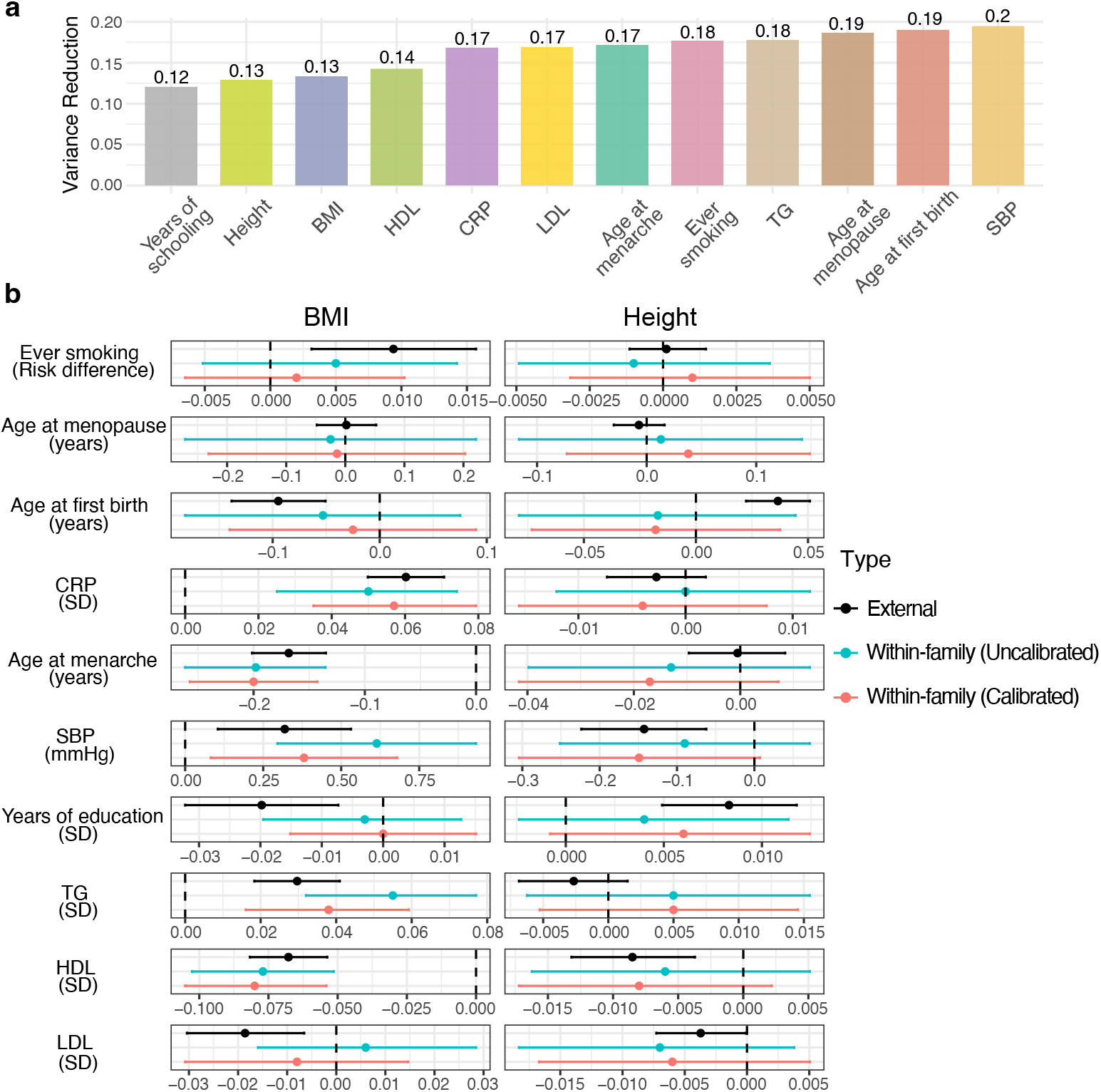
Calibration applied to published within-sibship GWAS summary statistics. a) Estimated variance reduction for within-family SNP–trait association estimates after calibration (constant for all SNPs). b) MR estimates for BMI and height using three sets of summary statistics: population-based (external GWAS), within-family uncalibrated, and within-family calibrated. Point estimates are shown with 95% confidence intervals, computed using MR.RAPS.

We then performed MR using both IVW, as in the original study, and MR.RAPS, applied to external population-based GWAS and to within-family GWAS before and after calibration. As in the original Howe et al. analysis, many MR effect estimates using population-based GWAS attenuated or became insignificant after family adjustment, including effects of height on age at first birth, SBP, HDL and years of education, and effects of BMI on age at first birth, years of education and smoking (Figure 4b). This pattern highlights the importance of controlling for genetic nurture, assortative mating, and shared environmental pathways.

Calibration further improved the precision of within-family MR. Confidence intervals narrowed for both IVW and MR.RAPS, while point estimates remained consistent with uncalibrated within-family results. Results using MR.RAPS are presented because it provides more stable inference under weak or noisy instruments [Wang et al., 2021]. Corresponding IVW results, which closely agree with those originally reported by Howe et al., are shown in Figure S2.

Taken together, these results demonstrate that our method integrates seamlessly with existing within-family summary statistics, enhances efficiency with negligible computational cost, and strengthens downstream causal inference without altering the unbiasedness inherent to within-family designs.

## 4 Discussion

Family-based GWAS provide unbiased estimates of direct genetic effects by leveraging random Mendelian segregation [Kong et al., 2018; Selzam et al., 2019; Warrington et al., 2018]. However, the limited availability of large family cohorts leads to substantially reduced statistical power compared with standard population GWAS, which constrains both discovery and downstream analyses such as Mendelian randomization. Our calibration framework offers a practical way to improve statistical efficiency by leveraging large-scale population-based GWAS while preserving the unbiasedness of within-family estimates.

Calibration may yield substantial efficiency gains in practice. Consistent with the theoretical predictions, within-family effect estimates showed variance reductions of up to 50% in trio designs, and up to 25% in sibling designs, depending on the sibling phenotypic correlation. These improvements correspond to an effective internal sample-size increase by a factor of 2 for trios and 1.33 for siblings. Recent work has explored related calibration ideas in other genetic contexts [Miao and Lu, 2024; Miao et al., 2024]. Our contribution is to develop a concrete and efficient calibration method tailored to family-based GWAS and only using the summary statistics.

A key benefit of our approach is that it does not require imputation of parental genotypes or access to individual-level data. This substantially reduces computational burden, avoids many of the logistical and privacy constraints associated with individual-level analyses, and makes the method applicable in settings where only published GWAS statistics are available. An equally important advantage is the flexibility of the calibration framework with respect to modeling choices. It does not rely on linearity assumptions and can be applied to summary statistics derived from logistic regression or other generalized linear models. This makes the method naturally suited for binary as well as continuous traits, an increasingly important consideration for large biobank studies.

Calibration improves precision at the SNP level, and these gains can propagate to downstream analyses such as MR. The extent to which MR benefits, however, depends on the strength of the instruments, the presence of pleiotropy and the MR method used, so reductions in GWAS-level variance do not always lead to proportionate reductions in MR uncertainty. Our results demonstrate that calibration can narrow confidence intervals for most traits, but the magnitude of improvement varies across datasets and phenotypes. Overall, the method strengthens within-family MR at essentially no additional computational cost while preserving the robustness of family-based inference.

One practical limitation is that the required internal summary statistics, namely the within-family association and the internal population-based association, are currently reported in only a small number of studies. In particular, internal population-based GWAS results are rarely released even when family-based analyses have been conducted. Wider adoption of reporting these statistics would make calibration feasible for a much broader range of datasets and would substantially increase the usefulness of within-family designs for downstream causal inference.

In summary, our calibration method provides a practical and scalable approach for increasing the efficiency of family-based GWAS and their downstream applications. By requiring only summary statistics and accommodating diverse modeling choices, it offers a general solution for improving precision while preserving the robustness of within-family designs.

## Data Availability

This study used controlled-access individual-level data from the UK Biobank, available through application to the UK Biobank. Publicly available summary statistics were obtained from OpenGWAS. The code used in this study is publicly available.

## 5 Data and code availability

UK Biobank individual-level participant data are available via requests to the UK Biobank access management team. The within-sibship summary statistics are available from the Within-Family GWAS Consortium on OpenGWAS (https://opengwas.io/). For each trait, we list the corresponding population-level code first, followed by the sibling-based code in parentheses: BMI (ieu-b-4816, ieu-b-4815), SBP (ieu-b-4818, ieu-b-4817), age at first birth (ieu-b-4820, ieu-b-4819), LDL (ieu-b-4846, ieu-b-4845), HDL (ieu-b-4844, ieu-b-4843), ever smoked (ieu-b-4858, ieu-b-4857), CRP (ieu-b-4764, ieu-b-4763), age at menarche (ieu-b-4822, ieu-b-4821), age at menopause (ieu-b-4824, ieu-b-4823), height (ieu-b-4814, ieu-b-4813), years of education (ieu-b-4836, ieu-b-4835), and TG (ieu-b-4850, ieu-b-4849).

The code for the proposed method can be accessed at (https://github.com/ZixuanWu1/CalibrationGWAS).

## 7 Acknowledgments

J.W. is partly supported by the National Science Foundation under grant DMS-2238656. M.S. is partly supported by the National Institutes of Health under grant HG002585. This research has been conducted using the UK Biobank Resource under application number 27386. We thank the University of Chicago Research Computing Center for computational support, and we are grateful to Peter Carbonetto for assistance with processing the UK Biobank data.

