## Supplemental Materials for "Improving estimation efficiencies for family-based GWAS by integrating large external data"

#### 1 Additional methodological details and derivations

##### 1.1 Equivalence of Common Within-Family Study Designs

This section provides the algebraic justification for the equivalence of the trio- and sibling-based specifications summarized in Table 1 of the main text. These derivations formalize why several widely used regression formulations target the same direct genetic effect.

###### 1.1.1 Trio designs

Consider the model that adjusts for the sum of maternal and paternal genotypes,

$$g\left(\mathbb{E}[Y_i \mid G_{ij}, G_{ij}^{pa}, \boldsymbol{C}_i]\right) = \tau_j G_{ij} + \delta_j G_{ij}^{pa} + \boldsymbol{\gamma}_j^\top \boldsymbol{C}_i.$$

where $G_{ij}^{pa} = G_{ij}^{f} + G_{ij}^{m}$. Because the parental genotype also decomposes as $G_{ij}^{pa} = T_{ij} + NT_{ij}$, with $T_{ij} = G_{ij}$ the transmitted allele and $NT_{ij}$ the non-transmitted allele, the model implies

$$g\left(\mathbb{E}[Y_i \mid G_{ij}, NT_{ij}, \boldsymbol{C}_i]\right) = (\tau_j + \delta_j) G_{ij} + \delta_j NT_{ij} + \boldsymbol{\gamma}_j^\top \boldsymbol{C}_i.$$

Thus, the direct effect of the transmitted allele equals the coefficient difference $\tau_j = \text{coef}(T_{ij}) - \text{coef}(NT_{ij})$ which is the estimand used when explicitly adjusting for non-transmitted alleles [1]. Therefore, adjusting for the sum of parental genotypes or non-transmitted alleles yields equivalent direct-effect estimators.

###### 1.1.2 Sibling designs

Let a family contain two siblings indexed by 1 and 2. Consider the model that adjusts for the total sibling genotype $F_{ij} = G_j^{sib} = G_{1j} + G_{2j}$ and uses a linear regression:

$$\mathbb{E}[Y_i \mid G_{ij}, (G_{i'j})_{i' \neq i}, \boldsymbol{C}_i] = \tau_j G_{ij} + \delta_j G_j^{sib} + \boldsymbol{\gamma}_j^\top \boldsymbol{C}_i.$$

Further, we assume additional covariates satisfy $Y_i \perp \boldsymbol{C}_{i^{\text{sib}}} \mid G_{ij}, G_{i^{\text{sib}}j}, \boldsymbol{C}_i$. Then, taking the difference between siblings gives

$$\mathbb{E}[Y_1 - Y_2 \mid G_{1j}, G_{2j}, \boldsymbol{C}_1, \boldsymbol{C}_2] = \tau_j (G_{1j} - G_{2j}) + \boldsymbol{\gamma}_j^\top (\boldsymbol{C}_1 - \boldsymbol{C}_2).$$

Thus, the regression coefficient on the genotype difference $G_{1j} - G_{2j}$ equals the direct genetic effect. Consequently, adjusting for the sibling genotype sum or fitting a difference regression when $K = 2$ targets the same estimand.

#### 1.2 Asymptotic statistical inference for the calibrated estimator

Here we provide full details of the joint distribution of $(\widehat{\tau}_j, \widehat{\alpha}_j^{\text{int}}, \widehat{\alpha}_j^{\text{ext}})$ and derive the asymptotic distribution of the calibrated estimator $\widehat{\tau}_j^{\text{cal}}$. As described in the Methods section, the within-family estimator $\widehat{\tau}_j$ is obtained by fitting a generalized linear model that conditions on family genotype:

$$g\left(\mathbb{E}[Y_i|G_{ij}, \boldsymbol{F}_{ij}, \boldsymbol{C}_i]\right) = \tau_j G_{ij} + \boldsymbol{\delta}_j^\top \boldsymbol{F}_{ij} + \boldsymbol{\gamma}_j^\top \boldsymbol{C}_i. \tag{1}$$

The internal and external population-based estimators $\widehat{\alpha}_j^{\text{int}}$ and $\widehat{\alpha}_j^{\text{ext}}$ are obtained by fitting the reduced model,

$$g\left(\mathbb{E}[Y_i|G_{ij}, \boldsymbol{C}_i]\right) = \alpha_j G_{ij} + \boldsymbol{\eta}_j^\top \boldsymbol{C}_i \tag{2}$$

We first state three key assumptions:

**Assumption 1.** *The full model in* (1) *is correctly specified for both the internal and external datasets.*

That is, the underlying data-generating process is assumed to follow the full model, while the reduced model in equation (2) may be misspecified. We further assume distributional homogeneity:

**Assumption 2.** *For any SNP $j$, the joint distribution of $(G_{ij}, \boldsymbol{F}_{ij}, \boldsymbol{C}_i, Y_i)$ is the same across individuals in both the internal and external datasets.*

Finally, we allow for weak dependence among individuals by assuming that each individual may be genetically related to at most a constant number of other individuals, even as the sample size grows:

**Assumption 3.** *For any individual $i$ in either the internal and external dataset, there exist at most $D = O(1)$ other individuals who are genetically dependent (e.g., related) with $i$, as the number of total individuals goes to infinity.*

Let $N_{\text{int}}$ and $N_{\text{ext}}$ denote the sample sizes of the internal and external datasets, respectively, with $N_{\text{int}} < N_{\text{ext}}$. Under these assumptions, we have the following result (proof in Section 4.1):

**Proposition 1.1.** *Under Assumptions 1-3,*

$$\lim_{N_{int}\to\infty} \mathbb{E}(\widehat{\alpha}_j^{int} - \widehat{\alpha}_j^{ext}) \to 0.$$

This implies a class of asymptotically unbiased calibrated estimators for $\tau_j$:

$$\widehat{\tau}_j^{\text{cal}}(\lambda) = \widehat{\tau}_j + \lambda(\widehat{\alpha}_j^{\text{int}} - \widehat{\alpha}_j^{\text{ext}}), \quad \lambda \in \mathbb{R},$$

and we can choose the optimal $\lambda_j^*$ for each SNP to minimize its asymptotic variance.

To derive the joint distribution of $(\widehat{\tau}_j, \widehat{\alpha}_j^{\text{int}}, \widehat{\alpha}_j^{\text{ext}})$, denote by $\widehat{\boldsymbol{b}}_j = (\widehat{\tau}_j, \widehat{\boldsymbol{\delta}}_j, \widehat{\boldsymbol{\gamma}}_j)$ the estimator for the full model parameter vector $\boldsymbol{b}_j = (\tau_j, \boldsymbol{\delta}_j, \boldsymbol{\gamma}_j)$, and $\widehat{\boldsymbol{a}}_j^{\text{int}} = (\widehat{\alpha}_j^{\text{int}}, \widehat{\boldsymbol{\eta}}_j^{\text{int}})$, and by $\widehat{\boldsymbol{a}}_j^{\text{ext}} = (\widehat{\alpha}_j^{\text{ext}}, \widehat{\boldsymbol{\eta}}_j^{\text{ext}})$ the estimators for the reduced model parameter vector $\boldsymbol{a}_j = (\alpha_j, \boldsymbol{\eta}_j)$ based on the internal and external data, respectively.

Define the score function for the full model as

$$\boldsymbol{s}_{ij}(\boldsymbol{b}_j) = \Big(Y_i - g^{-1}(\tau_j G_{ij} + \boldsymbol{\delta}_j^\top \boldsymbol{F}_{ij} + \boldsymbol{\gamma}_j^\top \boldsymbol{C}_i)\Big) \begin{pmatrix} G_{ij} \\ \boldsymbol{F}_{ij} \\ \boldsymbol{C}_{ij} \end{pmatrix}$$

and the score function for the reduced model as

$$\tilde{\boldsymbol{s}}_{ij}(\boldsymbol{a}_j) = \Big(Y_i - g^{-1}(\alpha_j G_{ij} + \boldsymbol{C}_i^\top \boldsymbol{\eta}_j)\Big) \begin{pmatrix} G_{ij} \\ \boldsymbol{C}_{ij} \end{pmatrix}.$$

Let $\boldsymbol{b}_j^*$ denote the true parameter value of $\boldsymbol{b}_j$ under Assumption 1, and define $\boldsymbol{a}_j^*$ as the solution to $\mathbb{E}\left[\tilde{\boldsymbol{s}}_{ij}(\boldsymbol{a}_j)\right] = \boldsymbol{0}$ assuming a unique solution exists. Under Assumption 2, the distribution of $(G_{ij}, \boldsymbol{F}_{ij}, \boldsymbol{C}_i)$ is the same across individuals, so this population moment is well-defined.

Let $\mathcal{D}_{\text{int}}$ and $\mathcal{D}_{\text{ext}}$ be the set of internal and external individuals, respectively. Then, $\widehat{\boldsymbol{b}}_j$ solves the score equation:

$$\sum_{i \in \mathcal{D}_{\text{int}}} \boldsymbol{s}_{ij}(\widehat{\boldsymbol{b}}_j) = \boldsymbol{0},$$

and $\widehat{\boldsymbol{a}}_j^{\text{int}}$ and $\widehat{\boldsymbol{a}}_j^{\text{ext}}$ solve the score equations

$$\sum_{i \in \mathcal{D}_{\text{int}}} \tilde{\boldsymbol{s}}_{ij}(\widehat{\boldsymbol{a}}_j^{\text{int}}) = \boldsymbol{0}, \quad \sum_{i \in \mathcal{D}_{\text{ext}}} \tilde{\boldsymbol{s}}_{ij}(\widehat{\boldsymbol{a}}_j^{\text{ext}}) = \boldsymbol{0}.$$

We can now characterize their joint asymptotic distribution (proof in Section 4.2).

**Proposition 1.2.** *Define*

$$\boldsymbol{D}_1 \equiv \mathbb{E}\left[\frac{\partial \boldsymbol{s}_{ij}(\boldsymbol{b}_j^*)}{\partial \boldsymbol{b}_j}\right], \quad \boldsymbol{D}_2 = \boldsymbol{D}_3 \equiv \mathbb{E}\left[\frac{\partial \tilde{\boldsymbol{s}}_{ij}(\boldsymbol{\alpha}_j^*)}{\partial \boldsymbol{a}_j}\right].$$

*Assume that* $\lim_{N_{int}\to\infty} \frac{N_{int}}{N_{ext}} \to \delta \geq 0$, *and that the following limiting covariances exist:*

$$\boldsymbol{C}_{11} = \lim_{N_{int}\to\infty} \frac{Cov\left(\sum_{i\in\mathcal{D}_{int}} \mathbf{s}_{ij}\left(\boldsymbol{b}_j^*\right)\right)}{N_{int}}, \quad \boldsymbol{C}_{22} = \lim_{N_{int}\to\infty} \frac{Cov\left(\sum_{i\in\mathcal{D}_{int}} \tilde{\mathbf{s}}_{ij}\left(\boldsymbol{\alpha}_j^*\right)\right)}{N_{int}}$$

$$\boldsymbol{C}_{33} = \lim_{N_{ext}\to\infty} \frac{Cov\left(\sum_{i\in\mathcal{D}_{ext}} \tilde{\mathbf{s}}_{ij}\left(\boldsymbol{\alpha}_j^*\right)\right)}{N_{ext}}$$

$$\boldsymbol{C}_{12} = \lim_{N_{int}\to\infty} \frac{Cov\left(\sum_{i\in\mathcal{D}_{int}} \mathbf{s}_{ij}\left(\boldsymbol{b}_j^*\right), \sum_{i\in\mathcal{D}_{int}} \tilde{\boldsymbol{s}}_{ij}\left(\boldsymbol{\alpha}_j^*\right)\right)}{N_{int}}$$

$$\boldsymbol{C}_{13} = \lim_{N_{int}\to\infty} \frac{Cov\left(\sum_{i\in\mathcal{D}_{int}} \mathbf{s}_{ij}\left(\boldsymbol{b}_j^*\right), \sum_{i\in\mathcal{D}_{ext}} \tilde{\boldsymbol{s}}_{ij}\left(\boldsymbol{\alpha}_j^*\right)\right)}{\sqrt{N_{int}N_{ext}}}$$

$$\boldsymbol{C}_{23} = \lim_{N_{int}\to\infty} \frac{Cov\left(\sum_{i\in\mathcal{D}_{int}} \tilde{\boldsymbol{s}}_{ij}\left(\boldsymbol{\alpha}_j^*\right), \sum_{i\in\mathcal{D}_{ext}} \tilde{\boldsymbol{s}}_{ij}\left(\boldsymbol{\alpha}_j^*\right)\right)}{\sqrt{N_{int}N_{ext}}}$$

*Then, under Assumptions 1-3 and additional regularity conditions (Assumption 10),*

$$\sqrt{N_{int}} \begin{pmatrix} \widehat{\boldsymbol{b}}_j - \boldsymbol{b}_j^* \\ \widehat{\boldsymbol{a}}_j^{int} - \boldsymbol{\alpha}_j^* \\ \widehat{\boldsymbol{a}}_j^{ext} - \boldsymbol{\alpha}_j^* \end{pmatrix} \to \mathcal{N}(\boldsymbol{0}, \boldsymbol{V}), \tag{3}$$

*where the asymptotic covariance matrix* $\boldsymbol{V}$ *has block form*

$$\boldsymbol{V} = \begin{pmatrix} \boldsymbol{D}_1^{-1}\boldsymbol{C}_{11}\boldsymbol{D}_1^{-1} & \boldsymbol{D}_1^{-1}\boldsymbol{C}_{12}\boldsymbol{D}_2^{-1} & \sqrt{\delta}\boldsymbol{D}_1^{-1}\boldsymbol{C}_{13}\boldsymbol{D}_3^{-1} \\ \boldsymbol{D}_2^{-1}\boldsymbol{C}_{12}^\top\boldsymbol{D}_1^{-1} & \boldsymbol{D}_2^{-1}\boldsymbol{C}_{22}\boldsymbol{D}_2^{-1} & \sqrt{\delta}\boldsymbol{D}_2^{-1}\boldsymbol{C}_{23}\boldsymbol{D}_3^{-1} \\ \sqrt{\delta}\boldsymbol{D}_3^{-1}\boldsymbol{C}_{13}^\top\boldsymbol{D}_1^{-1} & \sqrt{\delta}\boldsymbol{D}_3^{-1}\boldsymbol{C}_{23}^\top\boldsymbol{D}_2^{-1} & \delta\boldsymbol{D}_3^{-1}\boldsymbol{C}_{33}\boldsymbol{D}_3^{-1} \end{pmatrix}.$$

Based on Proposition 1.2, the components of $\boldsymbol{V}$ can be estimated from individual-level data using empirical means and covariances. In particular, the derivatives $\boldsymbol{D}_1$, $\boldsymbol{D}_2$ and $\boldsymbol{D}_3$ can be estimated by:

$$\widehat{\boldsymbol{D}}_1 = \frac{1}{N_{\text{int}}} \sum_{i \in \mathcal{D}_{\text{int}}} \frac{\partial \boldsymbol{s}_{ij}(\widehat{\boldsymbol{b}}_j)}{\partial \boldsymbol{b}_j}, \quad \widehat{\boldsymbol{D}}_2 = \frac{1}{N_{\text{int}}} \sum_{i \in \mathcal{D}_{\text{int}}} \frac{\partial \tilde{\boldsymbol{s}}_{ij}(\widehat{\boldsymbol{a}}_j^{\text{int}})}{\partial \boldsymbol{a}_j}, \quad \widehat{\boldsymbol{D}}_3 = \frac{1}{N_{\text{ext}}} \sum_{i \in \mathcal{D}_{\text{ext}}} \frac{\partial \tilde{\boldsymbol{s}}_{ij}(\widehat{\boldsymbol{a}}_j^{\text{ext}})}{\partial \boldsymbol{a}_j}.$$

To account for local dependence in estimating the covariance terms $\boldsymbol{C}_{kl}$, let $\mathcal{D}_i$ denote the set of individuals genetically related to $i$ (including $i$ itself), which we assume is known. Each covariance component $\boldsymbol{C}_{kl}$ can then be estimated via a double summation over dependent pairs:

$$\widehat{\boldsymbol{C}}_{11} = \frac{\sum_{i \in \mathcal{D}_{\text{int}}} \sum_{i' \in \mathcal{D}_i \cap \mathcal{D}_{\text{int}}} \mathbf{s}_{ij}(\widehat{\boldsymbol{b}}_j) \mathbf{s}_{i'j}(\widehat{\boldsymbol{b}}_j)^\top}{N_{\text{int}}}, \quad \widehat{\boldsymbol{C}}_{22} = \frac{\sum_{i \in \mathcal{D}_{\text{int}}} \sum_{i' \in \mathcal{D}_i \cap \mathcal{D}_{\text{int}}} \tilde{\boldsymbol{s}}_{ij}(\widehat{\boldsymbol{a}}_j^{\text{int}}) \tilde{\boldsymbol{s}}_{i'j}(\widehat{\boldsymbol{a}}_j^{\text{int}})^\top}{N_{\text{int}}},$$

$$\widehat{\boldsymbol{C}}_{33} = \frac{\sum_{i \in \mathcal{D}_{\text{ext}}} \sum_{i' \in \mathcal{D}_i \cap \mathcal{D}_{\text{ext}}} \tilde{\boldsymbol{s}}_{ij}(\widehat{\boldsymbol{a}}_j^{\text{ext}}) \tilde{\boldsymbol{s}}_{i'j}(\widehat{\boldsymbol{a}}_j^{\text{ext}})^\top}{N_{\text{ext}}}, \quad \widehat{\boldsymbol{C}}_{12} = \frac{\sum_{i \in \mathcal{D}_{\text{int}}} \sum_{i' \in \mathcal{D}_i \cap \mathcal{D}_{\text{int}}} \mathbf{s}_{ij}(\widehat{\boldsymbol{b}}_j) \tilde{\boldsymbol{s}}_{i'j}(\widehat{\boldsymbol{a}}_j^{\text{int}})^\top}{N_{\text{int}}},$$

$$\widehat{\boldsymbol{C}}_{13} = \frac{\sum_{i \in \mathcal{D}_{\text{int}}} \sum_{i' \in \mathcal{D}_i \cap \mathcal{D}_{\text{ext}}} \mathbf{s}_{ij}(\widehat{\boldsymbol{b}}_j) \tilde{\boldsymbol{s}}_{i'j}(\widehat{\boldsymbol{a}}_j^{\text{ext}})^\top}{\sqrt{N_{\text{int}} N_{\text{ext}}}}, \quad \widehat{\boldsymbol{C}}_{23} = \frac{\sum_{i \in \mathcal{D}_{\text{int}}} \sum_{i' \in \mathcal{D}_i \cap \mathcal{D}_{\text{ext}}} \tilde{\boldsymbol{s}}_{ij}(\widehat{\boldsymbol{a}}_j^{\text{int}}) \tilde{\boldsymbol{s}}_{i'j}(\widehat{\boldsymbol{a}}_j^{\text{ext}})^\top}{\sqrt{N_{\text{int}} N_{\text{ext}}}}.$$

To study the joint asymptotic distribution of $(\widehat{\tau}_j - \tau_j^*, \widehat{\alpha}_j^{\text{int}} - \widehat{\alpha}_j^{\text{ext}})$, note that this pair can be written as a linear transformation of the stacked parameter vector, i.e.,

$$\begin{pmatrix} \widehat{\tau}_j - \tau_j^* \\ \widehat{\alpha}_j^{\text{int}} - \widehat{\alpha}_j^{\text{ext}} \end{pmatrix} = \boldsymbol{A} \begin{pmatrix} \widehat{\boldsymbol{b}}_j - \boldsymbol{b}_j^* \\ \widehat{\boldsymbol{a}}_j^{\text{int}} - \boldsymbol{a}_j^* \\ \widehat{\boldsymbol{a}}_j^{\text{ext}} - \boldsymbol{a}_j^* \end{pmatrix}, \quad \boldsymbol{A} = \begin{pmatrix} 1 & \mathbf{0} & \mathbf{0} & 0 & \mathbf{0} & 0 & \mathbf{0} \\ 0 & \mathbf{0} & \mathbf{0} & 1 & \mathbf{0} & -1 & \mathbf{0} \end{pmatrix}.$$

Hence

$$\sqrt{N_{\text{int}}} \begin{pmatrix} \widehat{\tau}_j - \tau_j^* \\ \widehat{\alpha}_j^{\text{int}} - \widehat{\alpha}_j^{\text{ext}} \end{pmatrix} \to \mathcal{N}\left( \begin{pmatrix} 0 \\ 0 \end{pmatrix}, \boldsymbol{A}\boldsymbol{V}\boldsymbol{A}^\top \right).$$

Let $\frac{\boldsymbol{A}\widehat{\boldsymbol{V}}\boldsymbol{A}^\top}{N_{\text{int}}} = \begin{bmatrix} \widehat{v}_{11} & \widehat{v}_{12} \\ \widehat{v}_{21} & \widehat{v}_{22} \end{bmatrix}$. The optimal shrinkage coefficient is then

$$\widehat{\lambda}_j^* = -\widehat{v}_{12}/\widehat{v}_{22},$$

and the optimal calibrated estimator approximately follows a normal distribution:

$$\widehat{\tau}_j^{\text{cal}} = \widehat{\tau}_j + \widehat{\lambda}_j^* (\widehat{\alpha}_j^{\text{int}} - \widehat{\alpha}_j^{\text{ext}}) \approx \mathcal{N}\left( \tau_j, \widehat{v}_{11} - \frac{\widehat{v}_{12}^2}{\widehat{v}_{22}} \right).$$

This result enables construction of Wald-type confidence intervals and p-values for $\tau_j$, and guarantees that the calibrated estimator $\widehat{\tau}_j^{\text{cal}}$ has a smaller estimated standard error than the uncalibrated estimator $\widehat{\tau}_j$.

##### 1.3 Derivations of theoretical variance reduction

In this section, we provide theoretical approximation formulas for the variance reduction $\text{VR}_j$ under the weak single SNP effects.

##### 1.3.1 Approximation under weak single SNP effects

Our derivation relies on the assumption that each individual SNP has a weak effect on the traits of interest. Specifically, we assume:

**Assumption 4.** *For any SNP $j$, the direct and indirect SNP-trait associations are small in magnitude relative to the variance of the trait, i.e., $\tau_j^* << Var(Y_i)$ and $\boldsymbol{\delta}_j^* << Var(Y_i)$.*

Under this assumption, we should also have $\alpha_j^* << \mathrm{Var}(Y_i)$ and $\boldsymbol{\gamma}_j^* \approx \boldsymbol{\eta}_j^* \approx \boldsymbol{b}^*$ where $\boldsymbol{b}^*$ denotes the marginal regression coefficients of $Y_i$ on $\boldsymbol{C}_i$, defined as the solution to the moment condition:

$$\mathbb{E}\left[\left(Y_i - g^{-1}(\boldsymbol{C}_i^\top \boldsymbol{b}^*)\right)\boldsymbol{C}_i\right] = \mathbf{0}.$$

Let $\boldsymbol{X}_{ij} = (G_{ij}, \boldsymbol{F}_{ij})$ and assume without loss of generality that $\boldsymbol{X}_{ij}$ is centered at $\mathbf{0}$. We further assume that each $\boldsymbol{X}_{ij}$ for a single SNP $j$ and $\boldsymbol{C}_i$ are also weakly dependent:

**Assumption 5.** *For any SNP $j$, and any related pair of individuals $i$ and $i'$ (including the case $i = i'$), the conditional moments satisfy:*

$$\mathbb{E}\left[\boldsymbol{X}_{ij}\boldsymbol{X}_{i'j}^\top \mid (\boldsymbol{C}_i, \boldsymbol{C}_{i'}, Y_i, Y_{i'})\right] \approx \mathbb{E}\left[\boldsymbol{X}_{ij}\boldsymbol{X}_{i'j}^\top\right], \quad \mathbb{E}\left[\boldsymbol{X}_{ij} \mid (\boldsymbol{C}_i, \boldsymbol{C}_{i'}, Y_i, Y_{i'})\right] \approx \mathbf{0}.$$

Intuitively, this assumption reflects the idea that the distribution of $\boldsymbol{X}_{ij}$ is only weakly influenced by either the outcome $Y_i$ or covariates $\boldsymbol{C}_i$, which is reasonable under the assumption that the SNP-trait associations are small.

These assumptions imply that covariance matrices $\boldsymbol{D}_1, \boldsymbol{D}_2$ and $\boldsymbol{D}_3$ take approximately block diagonal forms. Specifically, for SNP $j$, denote

$$\mathrm{Var}(G_{ij}) = \mathbb{E}(G_{ij}^2) = \sigma_{Gj}^2, \quad \mathrm{Cov}(\boldsymbol{X}_{ij}) = \mathbb{E}\left(\boldsymbol{X}_{ij}\boldsymbol{X}_{ij}^\top\right) = \boldsymbol{\Sigma}_{Gj}.$$

We can obtain

$$\begin{aligned}
\boldsymbol{D}_1 &\approx -\begin{pmatrix} \mathbb{E}\left[g'(\boldsymbol{C}_i^\top \boldsymbol{b}^*)\right]\boldsymbol{\Sigma}_{Gj} & \mathbf{0} \\ \mathbf{0} & \mathbb{E}\left[g'(\boldsymbol{C}_i^\top \boldsymbol{b}^*)\boldsymbol{C}_i\boldsymbol{C}_i^\top\right] \end{pmatrix} \triangleq \begin{pmatrix} \tilde{\boldsymbol{D}}_1 & \mathbf{0} \\ \mathbf{0} & \boldsymbol{D}^* \end{pmatrix}, \\
\boldsymbol{D}_2 = \boldsymbol{D}_3 &\approx -\begin{pmatrix} \mathbb{E}\left[g'(\boldsymbol{C}_i^\top \boldsymbol{b}^*)\right]\sigma_{Gj}^2 & \mathbf{0} \\ \mathbf{0} & \mathbb{E}\left[g'(\boldsymbol{C}_i^\top \boldsymbol{b}^*)\boldsymbol{C}_i\boldsymbol{C}_i^\top\right] \end{pmatrix} \triangleq \begin{pmatrix} \tilde{\boldsymbol{D}}_2 & \mathbf{0} \\ \mathbf{0} & \boldsymbol{D}^* \end{pmatrix}.
\end{aligned} \tag{4}$$

Additionally, the covariance terms $\boldsymbol{C}_{kl}$ are also approximately block diagonal. Specifically, for a pair of individuals $i$ and $i'$, define the following quantities:

$$\sigma_{ii'} = \mathbb{E}\left[(Y_i - g^{-1}(\boldsymbol{C}_i^\top \boldsymbol{b}^*))(Y_{i'} - g^{-1}(\boldsymbol{C}_{i'}^\top \boldsymbol{b}^*))\right], \quad \boldsymbol{S}_{ii'} = \mathbb{E}\left[(Y_i - g^{-1}(\boldsymbol{C}_i^\top \boldsymbol{b}^*))(Y_{i'} - g^{-1}(\boldsymbol{C}_{i'}^\top \boldsymbol{b}^*))\boldsymbol{C}_i\boldsymbol{C}_{i'}^\top\right].$$

For SNP $j$, denote $\boldsymbol{\Lambda}_j = \mathrm{diag}(\sigma_{Gj}, \sqrt{\mathrm{diag}(\mathrm{Cov}(\boldsymbol{F}_{ij}))})$. Then under Assumptions 4-5, we can approximate the covariance matrices $\boldsymbol{C}_{kl}$ in the formula of as follows (omitting limits for readability):

$$
\begin{aligned}
\boldsymbol{C}_{11} &\approx -\begin{pmatrix} \frac{1}{N_{\text{int}}}\sum_{i,i'\in\mathcal{D}_{\text{int}}}\sigma_{ii'}\mathbb{E}\left(\boldsymbol{X}_{ij}\boldsymbol{X}_{i'j}^{\top}\right) & \boldsymbol{0} \\ \boldsymbol{0} & \frac{1}{N_{\text{int}}}\sum_{i,i'\in\mathcal{D}_{\text{int}}}\boldsymbol{S}_{ii'} \end{pmatrix} \triangleq \begin{pmatrix} \tilde{\boldsymbol{C}}_{11} & \boldsymbol{0} \\ \boldsymbol{0} & \boldsymbol{C}^{*} \end{pmatrix}, \\
\boldsymbol{C}_{12} &\approx -\begin{pmatrix} \frac{1}{N_{\text{int}}}\sum_{i,i'\in\mathcal{D}_{\text{int}}}\sigma_{ii'}\mathbb{E}\left(\boldsymbol{X}_{ij}G_{i'j}\right) & \boldsymbol{0} \\ \boldsymbol{0} & \frac{1}{N_{\text{int}}}\sum_{i,i'\in\mathcal{D}_{\text{int}}}\boldsymbol{S}_{ii'} \end{pmatrix} \triangleq \begin{pmatrix} \tilde{\boldsymbol{C}}_{12} & \boldsymbol{0} \\ \boldsymbol{0} & \boldsymbol{C}^{*} \end{pmatrix}, \\
\boldsymbol{C}_{13} &\approx \begin{pmatrix} \frac{1}{\sqrt{N_{\text{int}}N_{\text{ext}}}}\sum_{i\in\mathcal{D}_{\text{int}},i'\in\mathcal{D}_{\text{ext}}}\sigma_{ii'}\mathbb{E}\left(\boldsymbol{X}_{ij}G_{i'j}\right) & \boldsymbol{0} \\ \boldsymbol{0} & \frac{1}{\sqrt{N_{\text{int}}N_{\text{ext}}}}\sum_{i\in\mathcal{D}_{\text{int}},i'\in\mathcal{D}_{\text{ext}}}\boldsymbol{S}_{ii'} \end{pmatrix} \triangleq \begin{pmatrix} \tilde{\boldsymbol{C}}_{13} & \boldsymbol{0} \\ \boldsymbol{0} & \boldsymbol{C}^{**} \end{pmatrix}, \\
\boldsymbol{C}_{22} &\approx \begin{pmatrix} \frac{1}{N_{\text{int}}}\sum_{i,i'\in\mathcal{D}_{\text{int}}}\sigma_{ii'}\mathbb{E}\left(G_{ij}G_{i'j}\right) & \boldsymbol{0} \\ \boldsymbol{0} & \frac{1}{N_{\text{int}}}\sum_{i,i'\in\mathcal{D}_{\text{int}}}\boldsymbol{S}_{ii'} \end{pmatrix} \triangleq \begin{pmatrix} \tilde{\boldsymbol{C}}_{22} & \boldsymbol{0} \\ \boldsymbol{0} & \boldsymbol{C}^{*} \end{pmatrix}, \\
\boldsymbol{C}_{33} &\approx \begin{pmatrix} \frac{1}{N_{\text{ext}}}\sum_{i,i'\in\mathcal{D}_{\text{ext}}}\sigma_{ii'}\mathbb{E}\left(G_{ij}G_{i'j}\right) & \boldsymbol{0} \\ \boldsymbol{0} & \frac{1}{N_{\text{ext}}}\sum_{i,i'\in\mathcal{D}_{\text{ext}}}\boldsymbol{S}_{ii'} \end{pmatrix} \triangleq \begin{pmatrix} \tilde{\boldsymbol{C}}_{33} & \boldsymbol{0} \\ \boldsymbol{0} & \boldsymbol{C}^{***} \end{pmatrix}, \\
\boldsymbol{C}_{23} &\approx \begin{pmatrix} \frac{1}{\sqrt{N_{\text{int}}N_{\text{ext}}}}\sum_{i\in\mathcal{D}_{\text{int}},i'\in\mathcal{D}_{\text{ext}}}\sigma_{ii'}\mathbb{E}\left(G_{ij}G_{i'j}\right) & \boldsymbol{0} \\ \boldsymbol{0} & \frac{1}{\sqrt{N_{\text{int}}N_{\text{ext}}}}\sum_{i\in\mathcal{D}_{\text{int}},i'\in\mathcal{D}_{\text{ext}}}\boldsymbol{S}_{ii'} \end{pmatrix} \triangleq \begin{pmatrix} \tilde{\boldsymbol{C}}_{23} & \boldsymbol{0} \\ \boldsymbol{0} & \boldsymbol{C}^{**} \end{pmatrix}.
\end{aligned}
\tag{5}
$$

Given the above expressions, all the $\boldsymbol{D}_k$ and $\boldsymbol{C}_{kl}$ matrices are approximately block diagonal. And as a result, the genotype-related coefficients such as $(\widehat{\tau}_j, \widehat{\boldsymbol{\delta}}_j, \widehat{\alpha}_j^{\text{int}}, \widehat{\alpha}_j^{\text{ext}})$ are approximately independent from the covariate-related coefficients, which can greatly simplify our calculations, so we can basically ignore that fact that we are adjusting for additional covariates when performing statistical inference. For additional calculation and technical details to get these expressions, see Section 4.3. We will base on these derivations to calculate theoretical variance reduction in the trio and sibling designs, respectively.

##### 1.3.2 Trio Designs

To approximate variance reduction for the trio data, we further make the following assumption on genotypes:

**Assumption 6.**

$$Var(G_{ij}) = Var(G_{ij}^{m}) = Var(G_{ij}^{f})$$

This assumption states that the mother, father, and child have the same genotype variance at SNP $j$. It holds under Hardy–Weinberg equilibrium and random mating, where all individuals share allele frequency $f_j$ and the genotype variance of a biallelic SNP is $2f_j(1-f_j)$. Although real data may deviate from these idealized conditions due to population structure or assortative mating, we adopt this assumption to obtain closed-form approximations. Our simulations and UK Biobank analyses show that the resulting variance reduction formulas remain accurate even when the assumption is not exactly met.

In addition, we treat only the offspring from each trio as the observed samples and assume that all offspring are mutually independent (i.e. $\mathcal{D}_i = i$):

**Assumption 7.** *There is no residual familial correlation across different offspring. Individuals in the external dataset are mutually independent.*

Assumption 7 implies that for $i \neq i'$, we have $\mathbb{E}\left(\boldsymbol{X}_{ij}\boldsymbol{X}_{i'j}^{\top}\right) = \boldsymbol{0}$. Therefore, given Equation (5), we have

$$\tilde{\boldsymbol{C}}_{11} = \sigma_{Y|C}^{2}\boldsymbol{\Sigma}_{Gj}, \quad \tilde{\boldsymbol{C}}_{12} = \sigma_{Y|C}^{2}\text{Cov}(\boldsymbol{X}_{ij}, G_{ij}), \quad \tilde{\boldsymbol{C}}_{13} = \frac{N_{\text{share}}}{\sqrt{N_{\text{int}}N_{\text{ext}}}}\tilde{\boldsymbol{C}}_{12}$$

where $\sigma^2_{Y|C} \overset{\Delta}{\equiv} \sigma_{ii} = \mathbb{E}\left[(Y_i - g^{-1}(\boldsymbol{C}_i^\top \boldsymbol{b}^*))^2\right]$. Similarly, we can also show that

$$\tilde{\boldsymbol{C}}_{22} = \tilde{\boldsymbol{C}}_{33} = \sigma^2_{Y|C}\sigma^2_{Gj}, \quad \tilde{\boldsymbol{C}}_{13} = \frac{N_{\text{share}}}{\sqrt{N_{\text{int}}N_{\text{ext}}}}\tilde{\boldsymbol{C}}_{22},$$

where $N_{\text{share}}$ denotes the number of overlapping individuals between internal and external datasets. Given these formulas, we can have an analytical expression for the asymptotic variance of $(\widehat{\tau}_j, \widehat{\alpha}_j^{\text{int}}, \widehat{\alpha}_j^{\text{ext}})$.

For instance, for the case where $F_{ij} = G_{ij}^{pa}$, under Assumption 6, we have

$$\Sigma_{G_j} = \sigma^2_{Gj}\begin{pmatrix} 1 & 1+r_j \\ 1+r_j & 2(1+r_j) \end{pmatrix}, \quad \text{Cov}(\boldsymbol{X}_{ij}, G_{ij}) = \sigma^2_{Gj}\begin{pmatrix} 1 \\ 1+r_j \end{pmatrix}$$

where $r_j = \text{Cor}(G_{ij}, NT_{ij})$. Then we can derive the formula:

$$\text{Cov}\left(\begin{pmatrix} \widehat{\tau}_j \\ \widehat{\alpha}_j^{\text{int}} \\ \widehat{\alpha}_j^{\text{ext}} \end{pmatrix}\right) \approx \frac{\sigma^2_{Y|C}}{N_{\text{int}}\mathbb{E}\left[g'(\boldsymbol{C}_i^T\boldsymbol{b}^*)\right]^2\sigma^2_{Gj}}\begin{pmatrix} \frac{2}{1-r_j} & 1 & \frac{N_{\text{share}}}{N_{\text{ext}}} \\ 1 & 1 & \frac{N_{\text{share}}}{N_{\text{ext}}} \\ \frac{N_{\text{share}}}{N_{\text{ext}}} & \frac{N_{\text{share}}}{N_{\text{ext}}} & \frac{N_{\text{int}}}{N_{\text{ext}}} \end{pmatrix} \tag{6}$$

from which we can obtain

$$\text{Cov}\left(\begin{pmatrix} \widehat{\tau}_j \\ \widehat{\alpha}_j^{\text{int}} - \widehat{\alpha}_j^{\text{ext}} \end{pmatrix}\right) \approx \frac{\sigma^2_{Y|C}}{N_{\text{int}}\mathbb{E}\left[g'(\boldsymbol{C}_i^T\boldsymbol{b}^*)\right]^2\sigma^2_{Gj}}\begin{pmatrix} \frac{2}{1-r_j} & 1 - \frac{N_{\text{share}}}{N_{\text{ext}}} \\ 1 - \frac{N_{\text{share}}}{N_{\text{ext}}} & 1 + \frac{N_{\text{int}}}{N_{\text{ext}}} - \frac{2N_{\text{share}}}{N_{\text{ext}}} \end{pmatrix}. \tag{7}$$

As we demonstrated in Section 1.2, we know

$$\lambda_j^\star = \text{Cov}\left(\begin{pmatrix} \widehat{\tau}_j \\ \widehat{\alpha}_j^{\text{int}} - \widehat{\alpha}_j^{\text{ext}} \end{pmatrix}\right)_{12} \Big/ \text{Cov}\left(\begin{pmatrix} \widehat{\tau}_j \\ \widehat{\alpha}_j^{\text{int}} - \widehat{\alpha}_j^{\text{ext}} \end{pmatrix}\right)_{22} \approx \frac{1 - \frac{N_{\text{share}}}{N}}{1 + \frac{N_{\text{int}}}{N_{\text{ext}}} - \frac{2n_{shared}}{N}}$$

And thus the variance of the calibrated estimator is

$$\text{Var}\left(\widehat{\tau}_j + \lambda_j^\star(\widehat{\alpha}_j^{\text{int}} - \widehat{\alpha}_j^{\text{ext}})\right) \approx \frac{\sigma^2_{Y|C}}{N_{\text{int}}\mathbb{E}\left[g'(\boldsymbol{C}_i^\top\boldsymbol{b}^*)\right]^2\sigma^2_{Gj}}\left(\frac{2}{1-r_j} - \frac{(1-\frac{N_{\text{share}}}{N_{\text{ext}}})^2}{1 + \frac{N_{\text{int}}}{N_{\text{ext}}} - \frac{2n_{shared}}{N_{\text{ext}}}}\right)$$

The variance reduction is

$$\text{VR}_j = \frac{\text{Var}(\widehat{\tau}_j) - \text{Var}\left(\widehat{\tau}_j + \lambda_j^\star(\widehat{\alpha}_j^{\text{int}} - \widehat{\alpha}_j^{\text{ext}})\right)}{\text{Var}(\widehat{\tau}_j)} \approx \frac{1-r_j}{2}\frac{(1-\frac{N_{\text{share}}}{N_{\text{ext}}})^2}{\left(1 + \frac{N_{\text{int}}}{N_{\text{ext}}} - \frac{2N_{\text{share}}}{N_{\text{ext}}}\right)},$$

which approaches 0.5 for any SNP and any trait when $r_j = 0$ and $N_{\text{ext}}/N_{\text{int}} \to \infty$.

Due to the model equivalence as discussed in Section 1.1.1, the calibration estimator has the same reduction when $F_{ij} = NT_{ij}$. For the model that adjust for $\boldsymbol{F}_{ij} = (G_{ij}^f, G_{ij}^m)$, we can show that the variance reduction of the calibrated estimator is the same. The proof is provided in Section 4.4.

##### 1.3.3 Sibling Data

Our theoretical derivation is based on the following assumptions on the dependence structure of the sibling data.:

**Assumption 8.** *Siblings data satisfy:*

1. *Individuals across families are mutually independent in the internal data.*

2. *Each family has exactly $K$ siblings in the internal data, where the siblings do not contain identical twins.*

3. *The phenotypic correlation of the outcome between any two siblings, after adjusting the additional covariates $\boldsymbol{C}_i$, is the same.*

4. *For any SNP $j$, the genetic correlation between any two siblings, Cov$(\boldsymbol{X}_{ij}, \boldsymbol{X}_{i^{\text{sib}}j})$, is the same across siblings.*

5. *Individuals are mutually independent in the external data.*

Under Assumption 8, we define $\rho_Y \triangleq \mathbb{E}\left[(Y_i - g^{-1}(\boldsymbol{C}_i^\top \boldsymbol{b}^*))(Y_{i^{\text{sib}}} - g^{-1}(\boldsymbol{C}_{i^{\text{sib}}}^\top \boldsymbol{b}^*))\right]/\sigma^2_{Y|C}$, then given Equation (5), we have

$$\tilde{C}_{11} = \sigma^2_{Y|C}\boldsymbol{\Sigma}_{Gj} + (K-1)\sigma^2_{Y|C}\rho_Y \text{Cov}(\boldsymbol{X}_{ij}, \boldsymbol{X}_{i^{\text{sib}}j}).$$

$$\tilde{\boldsymbol{C}}_{12} = \sigma^2_{Y|C}\text{Cov}(\boldsymbol{X}_{ij}, G_{ij}) + (K-1)\sigma^2_{Y|C}\rho_Y \text{Cov}(\boldsymbol{X}_{ij}, G_{i^{\text{sib}}j}), \quad \tilde{\boldsymbol{C}}_{13} = \frac{N_{\text{share}}}{\sqrt{N_{\text{int}}N_{\text{ext}}}}\tilde{\boldsymbol{C}}_{12}$$

where $N_{\text{share}}$ is the number of shared individuals, thus also the number of shared families, where only one of the siblings in the family is assumed shared between the external and internal datasets.

Similarly, we also have

$$\tilde{\boldsymbol{C}}_{22} = \sigma^2_{Y|C}\sigma^2_{Gj} + (K-1)\sigma^2_{Y|C}\rho_Y \text{Cov}(G_{ij}, G_{i^{\text{sib}}j}), \quad \tilde{\boldsymbol{C}}_{33} = \sigma^2_{Y|C}\sigma^2_{Gj}, \quad \tilde{\boldsymbol{C}}_{23} = \frac{N_{\text{share}}}{\sqrt{N_{\text{int}}N_{\text{ext}}}}\tilde{\boldsymbol{C}}_{22}.$$

The family genotype is defined as $F_{ij} = \sum_{i' \in \mathcal{D}_i} G_{i'j}$, the summation of the genotype across all siblings in the same family. Denote $\kappa_j = \text{Cor}(G_{ij}, G_{i^{\text{sib}}j})$ as the genetic correlation between any two siblings, then we have

$$\Sigma_{Gj} = \text{Cov}(\boldsymbol{X}_{ij}, \boldsymbol{X}_{ij}) = \sigma^2_{Gj}\begin{pmatrix} 1 & 1+(K-1)\kappa_j \\ 1+(K-1)\kappa_j & K + K(K-1)\kappa_j \end{pmatrix},$$

$$\text{Cov}(\boldsymbol{X}_{ij}, \boldsymbol{X}_{i^{\text{sib}}j}) = \sigma^2_{Gj}\begin{pmatrix} \kappa_j & 1+(K-1)\kappa_j \\ 1+(K-1)\kappa_j & K + K(K-1)\kappa_j \end{pmatrix}.$$

Then we can derive

$$\begin{aligned} &\text{Cov}\left(\begin{pmatrix} \widehat{\tau}_j \\ \widehat{\alpha}_j^{\text{int}} \\ \widehat{\alpha}_j^{\text{ext}} \end{pmatrix}\right) \\ &\approx \frac{\sigma^2_{Y|C}}{N_{\text{int}}\mathbb{E}\left[g'(\boldsymbol{C}_i^\top \boldsymbol{b}^*)\right]^2 \sigma^2_{Gj}} \begin{pmatrix} \frac{K}{(K-1)(1-\kappa_j)}\left(1 - \frac{\rho_Y}{K-1}\right) & 1-\rho_Y & \frac{N_{\text{share}}}{N_{\text{ext}}}(1-\rho_Y) \\ 1-\rho_Y & x_j & \frac{N_{\text{share}}}{N_{\text{ext}}}x_j \\ \frac{N_{\text{share}}}{N_{\text{ext}}}(1-\rho_Y) & \frac{N_{\text{share}}}{N_{\text{ext}}}x_j & \frac{N_{\text{int}}}{N_{\text{ext}}} \end{pmatrix} \end{aligned} \tag{8}$$

with $x_j = 1 + (K-1)\rho_Y\kappa_j$. This indicates that

$$\begin{aligned} &\text{Cov}\left(\begin{pmatrix} \widehat{\tau}_j \\ \widehat{\alpha}_j^{\text{int}} - \widehat{\alpha}_j^{\text{ext}} \end{pmatrix}\right) \\ &\approx \frac{\sigma^2_{Y|C}}{N_{\text{int}}\mathbb{E}\left[g'(\boldsymbol{C}_i^\top \boldsymbol{b}^*)\right]^2 \sigma^2_{Gj}} \begin{pmatrix} \frac{K}{(K-1)(1-\kappa_j)}\left(1 - \frac{\rho_Y}{K-1}\right) & \left(1 - \frac{N_{\text{share}}}{N_{\text{ext}}}\right)(1-\rho_Y) \\ \left(1 - \frac{N_{\text{share}}}{N_{\text{ext}}}\right)(1-\rho_Y) & x_j + \frac{N_{\text{int}}}{N_{\text{ext}}} - \frac{2N_{\text{share}}}{N_{\text{ext}}}x_j \end{pmatrix}. \end{aligned} \tag{9}$$

As we demonstrated in Section 1.2, we know

$$\lambda_j^\star = \mathrm{Cov}\left(\begin{pmatrix}\widehat{\tau}_j \\ \widehat{\alpha}_j^{\mathrm{int}} - \widehat{\alpha}_j^{\mathrm{ext}}\end{pmatrix}\right)_{12} \Big/ \mathrm{Cov}\left(\begin{pmatrix}\widehat{\tau}_j \\ \widehat{\alpha}_j^{\mathrm{int}} - \widehat{\alpha}_j^{\mathrm{ext}}\end{pmatrix}\right)_{22} = \frac{\left(1 - \frac{N_{\mathrm{share}}}{N}\right)(1-\rho_Y)}{x_j + \frac{N_{\mathrm{int}}}{N_{\mathrm{ext}}} - \frac{2N_{\mathrm{share}}}{N_{\mathrm{ext}}} x_j}$$

and the variance of the calibrated estimator is

$$\mathrm{Var}\left(\widehat{\tau}_j + \lambda_j^\star(\widehat{\alpha}_j^{\mathrm{int}} - \widehat{\alpha}_j^{\mathrm{ext}})\right) = \frac{\sigma^2_{Y|C}}{N_{\mathrm{int}}\mathbb{E}\left[g'(\boldsymbol{C}_i^\top \boldsymbol{b}^*)\right]^2 \sigma^2_{Gj}} \left(\frac{K\left(1 - \frac{\rho_Y}{K-1}\right)}{(K-1)(1-\kappa_j)} - \frac{\left(1 - \frac{N_{\mathrm{share}}}{N_{\mathrm{ext}}}\right)^2 (1-\rho_Y)^2}{x_j + \frac{N_{\mathrm{int}}}{N_{\mathrm{ext}}} - \frac{2N_{\mathrm{share}}}{N_{\mathrm{ext}}} x_j}\right).$$

The variance reduction is

$$\mathrm{VR}_j = \frac{\mathrm{Var}(\widehat{\tau}_j) - \mathrm{Var}\left(\widehat{\tau}_j + \lambda_j^\star(\widehat{\alpha}_j^{\mathrm{int}} - \widehat{\alpha}_j^{\mathrm{ext}})\right)}{\mathrm{Var}(\widehat{\tau}_j)} \approx \frac{(K-1)(1-\kappa_j)}{K\left(1 - \frac{\rho_Y}{K-1}\right)} \frac{\left(1 - \frac{N_{\mathrm{share}}}{N_{\mathrm{ext}}}\right)^2 (1-\rho_Y)^2}{x_j + \frac{N_{\mathrm{int}}}{N_{\mathrm{ext}}} - \frac{2N_{\mathrm{share}}}{N_{\mathrm{ext}}} x_j}.$$

In particular, when $K = 2$, then $x_j = 1 + \rho_Y \kappa_j$, and the variance reduction formula can be simplified to:

$$\mathrm{VR}_j \approx \frac{1-\kappa_j}{2} \frac{\left(1 - \frac{N_{\mathrm{share}}}{N_{\mathrm{ext}}}\right)^2 (1-\rho_Y)}{\frac{N_{\mathrm{int}}}{N_{\mathrm{ext}}} + (1+\rho_Y\kappa_j)\left(1 - \frac{2N_{\mathrm{share}}}{N_{\mathrm{ext}}}\right)}.$$

As a special case, if $\kappa_j = 0.5$ (random mating) and $N_{\mathrm{ext}}/N_{\mathrm{int}} \to \infty$, the variance reduction can be further simplified to:

$$\frac{1-\rho_Y}{4 + 2\rho_Y},$$

which maximize at 1 when $\rho_Y = -1$, and minimize at 0 when $\rho_Y = 1$.

#### 1.4 Calibration only using summary statistics

To enable the implementation of the calibrated estimators and their variances using only GWAS summary statistics, we assume, specifically for each SNP $j$, access to the estimates $\widehat{\tau}_j, \widehat{\alpha}_j^{\mathrm{int}}$ and $\widehat{\alpha}_j^{\mathrm{ext}}$, along with their standard deviations. To calculate the calibrated estimators, our goal is to approximate the joint distribution of $(\widehat{\tau}_j - \tau_j^*, \widehat{\alpha}_j^{\mathrm{int}} - \widehat{\alpha}_j^{\mathrm{ext}})$ only using these summary-level information.

##### 1.4.1 Assuming shared mating pattern across SNPs

To simplify the joint covariance structure of $(\widehat{\tau}_j - \tau_j^*, \widehat{\alpha}_j^{\mathrm{int}} - \widehat{\alpha}_j^{\mathrm{ext}})$ so that there are shared components across SNPs, we may introduce an additional structural assumption on the correlation patterns of genotypes. Specifically, we assume that the correlation structure of genotypes between related individuals is invariant across SNPs. This reflects the idea that relatedness is determined by pedigree or shared ancestry, rather than by locus-specific features of individual SNPs.

**Assumption 9.** *For any two individuals $i$ and $i'$ (including the case $i = i'$), the genotype correlation:*

$$Cor\left(\boldsymbol{X}_{ij}, \boldsymbol{X}_{i'j}\right) = Cor\left(\begin{pmatrix} G_{ij} \\ \boldsymbol{F}_{ij}\end{pmatrix}, \begin{pmatrix} G_{i'j} \\ \boldsymbol{F}_{i'j}\end{pmatrix}\right) = \boldsymbol{\Phi}_{ii'}.$$

*is shared across all SNPs. We also define*

$$Cor\left(\boldsymbol{X}_{ij}, G_{i'j}\right) = \boldsymbol{\varphi}_{ii'}, \quad Cor\left(G_{ij}, G_{i'j}\right) = \phi_{ii'}.$$

This assumption is plausible in populations with stable mating structures, including random mating or assortative mating based on complex traits (e.g., height, education) that are influenced by many loci. Since mating is not based on genotypes at specific SNPs, the resulting patterns of relatedness affect the genome uniformly, inducing correlation structures that do not vary systematically across loci. With such a simplification, we can borrow information across SNPs to perform calibration only using GWAS summary statistics.

Specifically, for each SNP $j$, denote $\mathbf{\Lambda}_j = \text{diag}(\sigma_{Gj}, \sqrt{\text{diag}(\text{Cov}(\boldsymbol{F}_{ij}))})$. Then under Assumption 9, the $\tilde{\boldsymbol{C}}_{kl}$ as defined in Equation (5) can be simplified to (omitting limits for readability):

$$\tilde{\boldsymbol{C}}_{11} = \mathbf{\Lambda} \left( \frac{1}{N_{\text{int}}} \sum_{i,i' \in \mathcal{D}_{\text{int}}} \sigma_{ii'} \mathbf{\Phi}_{ii'} \right) \mathbf{\Lambda}, \quad \tilde{\boldsymbol{C}}_{12} = \mathbf{\Lambda} \left( \frac{1}{N_{\text{int}}} \sum_{i,i' \in \mathcal{D}_{\text{int}}} \sigma_{ii'} \boldsymbol{\varphi}_{ii'} \right) \sigma_{Gj},$$

$$\tilde{\boldsymbol{C}}_{13} = \mathbf{\Lambda} \left( \frac{1}{\sqrt{N_{\text{int}} N_{\text{ext}}}} \sum_{i \in \mathcal{D}_{\text{int}}, i' \in \mathcal{D}_{\text{ext}}} \sigma_{ii'} \boldsymbol{\varphi}_{ii'} \right) \sigma_{Gj}, \quad \tilde{\boldsymbol{C}}_{22} = \sigma_{Gj}^2 \cdot \frac{1}{N_{\text{int}}} \sum_{i,i' \in \mathcal{D}_{\text{int}}} \sigma_{ii'} \phi_{ii'},$$

$$\tilde{\boldsymbol{C}}_{33} = \sigma_{Gj}^2 \cdot \frac{1}{N_{\text{ext}}} \sum_{i,i' \in \mathcal{D}_{\text{ext}}} \sigma_{ii'} \phi_{ii'}, \quad \tilde{\boldsymbol{C}}_{23} = \sigma_{Gj}^2 \cdot \frac{1}{\sqrt{N_{\text{int}} N_{\text{ext}}}} \sum_{i \in \mathcal{D}_{\text{int}}, i' \in \mathcal{D}_{\text{ext}}} \sigma_{ii'} \phi_{ii'}.$$

Additionally,

$$\tilde{\boldsymbol{D}}_2 = \mathbb{E}\left[ g'(\boldsymbol{C}_i^\top \boldsymbol{b}^*) \right] \mathbf{\Lambda} \mathbf{\Phi}_{ii} \mathbf{\Lambda}$$

as under Assumption 2, the genotype correlation $\mathbf{\Phi}_{ii}$ is a constant across $i$. Then, combining with Equations (4) and (5), we have

$$\text{Cov}\left( \begin{pmatrix} \widehat{\alpha}_j^{\text{int}} \\ \widehat{\alpha}_j^{\text{ext}} \end{pmatrix} \right) \approx \begin{pmatrix} \tilde{\boldsymbol{D}}_2^{-1} \tilde{\boldsymbol{C}}_{22} \tilde{\boldsymbol{D}}_2^{-1} & \sqrt{\delta} \tilde{\boldsymbol{D}}_2^{-1} \tilde{\boldsymbol{C}}_{23} \tilde{\boldsymbol{D}}_2^{-1} \\ \sqrt{\delta} \tilde{\boldsymbol{D}}_2^{-1} \tilde{\boldsymbol{C}}_{23}^\top \tilde{\boldsymbol{D}}_2^{-1} & \delta \tilde{\boldsymbol{D}}_2^{-1} \tilde{\boldsymbol{C}}_{33} \tilde{\boldsymbol{D}}_2^{-1} \end{pmatrix}$$

$$= \frac{1}{N_{\text{int}} \mathbb{E}\left[ g'(\boldsymbol{C}_i^\top \boldsymbol{b}^*) \right]^2 \sigma_{Gj}^2} \begin{pmatrix} \frac{1}{N_{\text{int}}} \sum_{i,i' \in \mathcal{D}_{\text{int}}} \sigma_{ii'} \phi_{ii'} & \frac{1}{N_{\text{ext}}} \sum_{i \in \mathcal{D}_{\text{int}}, i' \in \mathcal{D}_{\text{ext}}} \sigma_{ii'} \phi_{ii'} \\ \frac{1}{N_{\text{ext}}} \sum_{i \in \mathcal{D}_{\text{int}}, i' \in \mathcal{D}_{\text{ext}}} \sigma_{ii'} \phi_{ii'} & \frac{N_{\text{int}}}{N_{\text{ext}}^2} \sum_{i,i' \in \mathcal{D}_{\text{ext}}} \sigma_{ii'} \phi_{ii'} \end{pmatrix}.$$

Thus,

$$\text{Cor}\left( \widehat{\alpha}_j^{\text{int}}, \widehat{\alpha}_j^{\text{ext}} \right) \approx \text{const.}, \quad \frac{\text{Var}\left( \widehat{\alpha}_j^{\text{int}} \right)}{\text{Var}\left( \widehat{\alpha}_j^{\text{ext}} \right)} \approx \text{const.},$$

meaning that both the correlation and relative scale of the internal and external population GWAS summary statistics are approximately shared across SNPs. Given this shared correlation and the marginal variances already provided in GWAS summary statistics, we can also estimate their differences' standard deviations.

Similarly, we can obtain

$$\text{Cor}\left( \widehat{\tau}_j, \widehat{\alpha}_j^{\text{int}} \right) \approx \text{const.}, \quad \text{Cor}\left( \widehat{\tau}_j, \widehat{\alpha}_j^{\text{ext}} \right) \approx \text{const.}.$$

which implies

$$\text{Cor}\left( \widehat{\tau}_j, \widehat{\alpha}_j^{\text{int}} - \widehat{\alpha}_j^{\text{ext}} \right) \approx \text{const.}$$

Since $\mathbb{E}(\widehat{\alpha}_j^{\text{int}} - \widehat{\alpha}_j^{\text{ext}}) \to 0$ as $n \to \infty$, we can empirically estimate this correlation across SNPs:

$$\widehat{\rho} = \widehat{\text{Cor}}\left( \widehat{\tau}_j, \widehat{\alpha}_j^{\text{int}} - \widehat{\alpha}_j^{\text{ext}} \right) = \frac{1}{J} \sum_{j=1}^{J} \frac{\widehat{\tau}_j (\widehat{\alpha}_j^{\text{int}} - \widehat{\alpha}_j^{\text{ext}})}{\widehat{\text{sd}}(\widehat{\tau}_j) \widehat{\text{sd}}(\widehat{\alpha}_j^{\text{int}} - \widehat{\alpha}_j^{\text{ext}})}$$

where $J$ is the total number of SNPs. This leads to a practical form of the calibrated estimator:

$$\widehat{\lambda}_j^* = -\widehat{\rho}\frac{\widehat{\mathrm{sd}}(\widehat{\tau}_j)}{\widehat{\mathrm{sd}}(\widehat{\alpha}_j^{\mathrm{int}} - \widehat{\alpha}_j^{\mathrm{ext}})}, \quad \widehat{\tau}_j^{\mathrm{cal}} = \widehat{\tau}_j + \widehat{\lambda}^*(\widehat{\alpha}_j^{\mathrm{int}} - \widehat{\alpha}_{1j}') \approx \mathcal{N}\left(\tau_j, (1-\widehat{\rho}^2)\widehat{\mathrm{sd}}(\widehat{\tau}_j)^2\right).$$

This representation highlights how shared correlation structures induced by mating patterns enable calibration with improved statistical efficiency using only summary-level data.

##### 1.4.2 Accounting for heterogeneous mating patterns across SNPs

We still construct the calibration estimator using summary statistics without Assumption 9, given that we have pre-computed the genotype correlations $\{\widehat{r}_j, j = 1, \ldots, J\}$, the correlations between transmitted and non-transmitted alleles for trio data, and $\{\widehat{\kappa}_j, j = 1, \ldots, J\}$, the correlations between the genotype of two siblings for sibling data.

**Trio data.** For trio data, under the assumptions of Section 1.3.2, even without Assumption 9, we still have Equations (6)-(7), which indicates that

$$\mathrm{Cor}\left(\widehat{\alpha}_j^{\mathrm{int}}, \widehat{\alpha}_j^{\mathrm{ext}}\right) \approx \frac{N_{\mathrm{share}}}{\sqrt{N_{\mathrm{int}}N_{\mathrm{ext}}}},$$

$$\rho_j = \mathrm{Cor}\left(\widehat{\tau}_j, \widehat{\alpha}_j^{\mathrm{int}} - \widehat{\alpha}_j^{\mathrm{ext}}\right) \approx \frac{N_{\mathrm{ext}} - N_{\mathrm{share}}}{\sqrt{N_{\mathrm{ext}}(N_{\mathrm{ext}} + N_{\mathrm{int}} - 2N_{\mathrm{share}})}}\sqrt{\frac{1-r_j}{2}}.$$

To estimate each $\rho_j$, we plug in pre-calculated estimates $\widehat{r}_j$ and estimate $\rho_j$ using the formula:

$$\widehat{\rho}_j = \widehat{\Delta}\sqrt{\frac{1-\widehat{r}_j}{2}}, \quad \widehat{\Delta} = \frac{1}{J}\sum_{j=1}^{J}\frac{\widehat{\tau}_j(\widehat{\alpha}_j^{\mathrm{int}} - \widehat{\alpha}_j^{\mathrm{ext}})}{\widehat{\mathrm{sd}}(\widehat{\tau}_j)\widehat{\mathrm{sd}}(\widehat{\alpha}_j^{\mathrm{int}} - \widehat{\alpha}_j^{\mathrm{ext}})}\sqrt{\frac{2}{1-\widehat{r}_j}}.$$

**Sibling data.** For sibling data, under the assumptions in 1.3.3, even without Assumption 9, we still have Equation (8)-(9), which indicates that

$$\eta_j = \mathrm{Cor}\left(\widehat{\alpha}_j^{\mathrm{int}}, \widehat{\alpha}_j^{\mathrm{ext}}\right) \approx \frac{N_{\mathrm{share}}}{\sqrt{N_{\mathrm{int}}N_{\mathrm{ext}}}}\sqrt{1 + (K-1)\rho_Y\kappa_j} \tag{10}$$

$$\rho_j = \mathrm{Cor}\left(\widehat{\tau}_j, \widehat{\alpha}_j^{\mathrm{int}} - \widehat{\alpha}_j^{\mathrm{ext}}\right) \approx \frac{N_{\mathrm{ext}} - N_{\mathrm{share}}}{\sqrt{N_{\mathrm{ext}}((N_{\mathrm{ext}} - 2N_{\mathrm{share}})x_j + N_{\mathrm{int}})}}\frac{1-\rho_Y}{\sqrt{1-\frac{\rho_Y}{K-1}}}\sqrt{\frac{(K-1)(1-\kappa_j)}{K}}. \tag{11}$$

If $K = 2$, we can simplify the formula to:

$$\eta_j = \mathrm{Cor}\left(\widehat{\alpha}_j^{\mathrm{int}}, \widehat{\alpha}_j^{\mathrm{ext}}\right) \approx \frac{N_{\mathrm{share}}}{\sqrt{N_{\mathrm{int}}N_{\mathrm{ext}}}}\sqrt{1 + \rho_Y\kappa_j}$$

$$\rho_j = \mathrm{Cor}\left(\widehat{\tau}_j, \widehat{\alpha}_j^{\mathrm{int}} - \widehat{\alpha}_j^{\mathrm{ext}}\right) \approx \frac{N_{\mathrm{ext}} - N_{\mathrm{share}}}{\sqrt{N_{\mathrm{ext}}((N_{\mathrm{ext}} - 2N_{\mathrm{share}})(1+\rho_Y\kappa_j) + N_{\mathrm{int}})}}\sqrt{1-\rho_Y}\sqrt{\frac{1-\kappa_j}{2}}.$$

To estimate each $\rho_j$, we first select a subset of SNPs $S = \{j : \widehat{\kappa}_j \approx 0.5\}$, and compute

$$\widehat{\eta}_{0.5} = \frac{1}{|S|}\sum_{j\in S}\frac{\widehat{\alpha}_j^{\mathrm{int}}\widehat{\alpha}_j^{\mathrm{ext}}}{\widehat{\mathrm{sd}}(\widehat{\alpha}_j^{\mathrm{int}})\widehat{\mathrm{sd}}(\widehat{\alpha}_j^{\mathrm{ext}})}, \qquad \widehat{\rho}_{0.5} = \frac{1}{|S|}\sum_{j\in S}\frac{\widehat{\tau}_j\left(\widehat{\alpha}_j^{\mathrm{int}} - \widehat{\alpha}_j^{\mathrm{ext}}\right)}{\widehat{\mathrm{sd}}(\widehat{\tau}_j)\widehat{\mathrm{sd}}(\widehat{\alpha}_j^{\mathrm{int}} - \widehat{\alpha}_j^{\mathrm{ext}})}.$$

We treat $\widehat{\eta}_{0.5}$ and $\widehat{\rho}_{0.5}$ as initial estimates of the correlations and substitute $\widehat{\eta}_{0.5}$, $\widehat{\rho}_{0.5}$, and $\kappa_j = 0.5$ into Equations (10)–(11), yielding

$$\widehat{\eta}_{0.5} = \frac{N_{\text{share}}}{\sqrt{N_{\text{int}} N_{\text{ext}}}} \sqrt{1 + \frac{(K-1)\rho_Y}{2}}, \tag{12}$$

$$\widehat{\rho}_{0.5} = \frac{N_{\text{ext}} - N_{\text{share}}}{\sqrt{N_{\text{ext}}\left((N_{\text{ext}} - 2N_{\text{share}})\left(1 + \frac{(K-1)\rho_Y}{2}\right) + N_{\text{int}}\right)}} \frac{1 - \rho_Y}{\sqrt{1 - \frac{\rho_Y}{K-1}}} \sqrt{\frac{K-1}{2K}}. \tag{13}$$

We solve this system of two equations to obtain estimates of $N_{\text{share}}$ and $\rho_Y$, which are then substituted back into Equations (10)–(11) to compute $\widehat{\eta}_j$ and $\widehat{\rho}_j$ for each SNP.

### 2 Additional simulation setup details

This section provides the full mathematical specification of the simulation model. A descriptive overview of the simulation design, including trio/sibling structures, allele-frequency sampling, and sample-size choices, is provided in the main text.

**Genotype model** The conceptual genotype-generation procedure is described in the main text. Here we record the formal model:

$$\boldsymbol{G}_i^f,\ \boldsymbol{G}_i^m \sim \text{Bin}(2, \boldsymbol{f}),$$

where $\boldsymbol{f}$ is the vector of SNP-specific allele frequencies. Offspring genotypes $\boldsymbol{G}_i$ follow Mendelian segregation, obtained by independently sampling one allele from each parent at every locus. When two offspring are required, this step is repeated independently.

**Parental phenotype model** The main text outlines the role of parental phenotypes in inducing indirect genetic effects. The exact generative model is:

$$\begin{aligned}
\boldsymbol{b}^{pa} &\sim N_p(\boldsymbol{0}, \sigma^2 I), \\
Y_i^f &= (\boldsymbol{G}_i^f)^\top \boldsymbol{b}^{pa} + \epsilon_i^f, \qquad \epsilon_i^f \sim N(0, \sigma^2), \\
Y_i^m &= (\boldsymbol{G}_i^m)^\top \boldsymbol{b}^{pa} + \epsilon_i^m, \qquad \epsilon_i^m \sim N(0, \sigma^2).
\end{aligned}$$

The vector $\boldsymbol{b}^{pa}$ is generated once and held fixed across simulation replicates.

**Offspring phenotype model** The main text describes how offspring traits incorporate direct and indirect effects. The precise continuous-trait model is:

$$\begin{aligned}
\boldsymbol{b} &\sim N_p(\boldsymbol{0}, \sigma^2 I), \\
Y_i &= \boldsymbol{G}_i^\top \boldsymbol{b} + r\,(Y_i^m + Y_i^f) + \epsilon_i, \qquad \epsilon_i \sim N(0, \sigma^2),
\end{aligned}$$

with $\boldsymbol{b}$ fixed across replicates. Binary traits are generated by applying a logistic link to the same linear predictor:

$$Y_i \sim \text{Bernoulli}\Big(h\Big(\boldsymbol{G}_i^\top \boldsymbol{b} + r(Y_i^m + Y_i^f) + \epsilon_i\Big)\Big), \qquad \epsilon_i \sim N(0, \sigma^2), \qquad h(x) = \frac{1}{1 + e^{-x}}.$$

**Simulation parameters** All sample size specifications and the number of simulation replicates are reported in the main text. In the trio setting, we set $\sigma = 3$ and vary the indirect-effect parameter $r \in \{0, 0.1, 0.2, 0.3, 0.4, 0.5\}$. In the sibling setting, we fix $r = 0.1$ and vary the residual variance using $\sigma \in \{2, 4, 6, 8, 10, 12\}$.

### 3 Additional Information for the within-sibship analysis

In the real-data analysis based on summary statistics, we rescale the external reference datasets to match the Within-Sibship Consortium internal population estimates using Mendelian randomization. Below in Table 1, we list the consortia used for this scale adjustment, including: (i) the selection datasets used to identify genome-wide significant SNPs (p-value $< 10^{-8}$); (ii) the external reference datasets.

| Trait | Selection source | Reference source |
|---|---|---|
| SBP | GERA [2] | Neale Lab |
| LDL | GERA [3] | Neale Lab |
| HDL | GERA [3] | Neale Lab |
| TG | GERA [3] | Neale Lab |
| BMI | GIANT | Neale Lab |
| Age at first birth | MRC-IEU | Neale Lab |
| Height | GIANT | Neale Lab |
| CRP | UK Biobank + CHARGE | Neale Lab |
| Age at Menopause | ReproGen | Neale Lab |
| Age at Menarche | ReproGen | MRC-IEU |
| Ever smoked | UK Biobank + TAG | MRC-IEU |
| Years of Schooling | MRC-IEU | SSGAC |

Table 1: Consortia, selection, and external sources.

The datasets used in this study can be obtained from the sources listed below. For traits available through OpenGWAS [4] (`https://opengwas.io/`) or Neale Lab (`https://www.nealelab.is/uk-biobank`), we provide their corresponding codes for convenience.

1. **GERA:** Available from the GWAS Catalog (`https://www.ebi.ac.uk/gwas/home`) with study codes: SBP (GCST007095), HDL (GCST007140), LDL (GCST007141), and TG (GCST007142).

2. **Neale Lab:** Downloadable from the Neale Lab UK Biobank repository (`https://www.nealelab.is/uk-biobank`) with trait codes: BMI (21001), age at first birth (2754), SBP (4080), LDL (30780), HDL (30760), TG (30870), age at menarche (2714), age at menopause (3581), ever smoked (20160), CRP (30710), and height (50).

3. **GIANT BMI:** Available at `https://portals.broadinstitute.org/collaboration/giant/index.php/GIANT_consortium_data_files#2017_Gene_x_Environment_Summary_Statistics`.

4. **Other (OpenGWAS):** Additional traits available through OpenGWAS (`https://opengwas.io`) with codes: age at menarche (MRC-IEU; ukb-b-3768), ever smoked (MRC-IEU; ukb-b-20261), years of education (SSGAC; ieu-a-1239), height (GIANT; ieu-a-89), age at menarche (ReproGen; ieu-a-1004), ever smoked (UK Biobank + TAG; ieu-a-89), CRP (UK Biobank + CHARGE; ieu-b-35), and age at first birth (MRC-IEU; ukb-b-12405).

### 4 Proofs and additional technical details

#### 4.1 Proof of Proposition 1.1

For the correctly-specified model (1), we denote $\boldsymbol{b}_j^* = (\tau_j^*, \boldsymbol{\delta}_j^*, \boldsymbol{\gamma}_j^*)$ as the true values of these parameters. For the mis-specified model (2), we define $\boldsymbol{\alpha}^* = (\alpha_j^*, \boldsymbol{\eta}_j^*)$ as the solution to

$$\begin{aligned}&\mathbb{E}\left[\left(Y_i - g^{-1}(\alpha_j G_{ij} + \boldsymbol{C}_i^\top \boldsymbol{\eta}_j)\right)\begin{pmatrix}G_{ij}\\ \boldsymbol{C}_i\end{pmatrix}\right]\\ =&\mathbb{E}\left[\left(g^{-1}(\tau_j^* G_{ij} + \boldsymbol{F}_{ij}^\top \boldsymbol{\delta}_j^* + \boldsymbol{C}_i^\top \boldsymbol{\gamma}_j^*) - g_2^{-1}(\alpha_j G_{ij} + \boldsymbol{C}_i^\top \boldsymbol{\eta}_j)\right)\begin{pmatrix}G_{ij}\\ \boldsymbol{C}_i\end{pmatrix}\right] = \mathbf{0}\end{aligned}$$

assuming that the solution is unique. Given Assumption 2, we can W.L.O.G. assume that for each SNP $j$, $(G_{ij}, \boldsymbol{F}_{ij}, \boldsymbol{C}_i)$ are identically distributed across $i$, indicating that $\boldsymbol{a}_j^*$ is well-defined.

Let $\widehat{\boldsymbol{a}}_j^{\text{int}}$ and $\widehat{\boldsymbol{a}}_j^{\text{ext}}$ be the solutions of the estimating equations of the reduced model (2) on internal and external data, respectively. For a sample $i$ either in the internal or external data, define

$$\tilde{\boldsymbol{s}}_{ij}(\boldsymbol{a}_j) = (Y_i - g^{-1}(\alpha_j G_{ij} + \boldsymbol{C}_i^\top \boldsymbol{\eta}_j))\begin{pmatrix}G_{ij}\\ \boldsymbol{C}_i\end{pmatrix},$$

then the estimators satisfy

$$\sum_{i\in\mathcal{D}_{\text{int}}} \tilde{\boldsymbol{s}}_{ij}(\widehat{\boldsymbol{a}}_j^{\text{int}}) = \mathbf{0}, \quad \sum_{\in\mathcal{D}_{\text{ext}}} \tilde{\boldsymbol{s}}_{ij}(\widehat{\boldsymbol{a}}_j^{\text{ext}}) = \mathbf{0}.$$

By the classical theory of estimating equations, under Assumption 3 we have

$$\mathbb{E}(\widehat{\boldsymbol{a}}_j^{\text{int}}) - \boldsymbol{a}_j^* \to 0, \quad \mathbb{E}(\widehat{\boldsymbol{a}}_j^{\text{ext}}) - \boldsymbol{a}_j^* \to 0$$

as $N_{\text{int}} \to \infty$.

#### 4.2 Proof of Proposition 1.2

Let $\boldsymbol{W}_{ij} = (G_{ij}, \boldsymbol{F}_{ij}, \boldsymbol{C}_{ij})$ and define

$$\boldsymbol{s}(\boldsymbol{W}_{ij}, \boldsymbol{b}) := \boldsymbol{s}_{ij}(\boldsymbol{b}) = (Y_i - \mu_{ij})\begin{pmatrix}G_{ij}\\ \boldsymbol{F}_{ij}\\ \boldsymbol{C}_{ij}\end{pmatrix}, \quad \tilde{\boldsymbol{s}}(\boldsymbol{W}_{ij}, \boldsymbol{\alpha}) := \tilde{\boldsymbol{s}}_{ij}(\boldsymbol{\alpha}) = (Y_i - \tilde{\mu}_{ij})\begin{pmatrix}G_{ij}\\ \boldsymbol{F}_{ij}\\ \boldsymbol{C}_{ij}\end{pmatrix}$$

Furthermore let

$$\boldsymbol{\theta}_j = \begin{pmatrix}\boldsymbol{b}_j\\ \boldsymbol{a}_j\end{pmatrix}, \quad \boldsymbol{u}(\boldsymbol{W}_{ij}, \boldsymbol{\theta}_j) = \begin{pmatrix}\boldsymbol{s}(\boldsymbol{W}_{ij}, \boldsymbol{b}_j)\\ \tilde{\boldsymbol{s}}(\boldsymbol{W}_{ij}, \boldsymbol{a}_j)\end{pmatrix}$$

**Assumption 10** (Regularity conditions)**.** *The regularity conditions required in proving the proposition listed below.*

1. $\boldsymbol{\theta}_j^\star = ((\boldsymbol{b}_j^\star)^\top, (\boldsymbol{\alpha}_j^\star)^\top)^\top$ *lies in the interior of a compact parameter space* $\boldsymbol{\Theta}$.

2. $\boldsymbol{u}$ *is continuous differentiable*

3. 
$$\mathbb{E}_{\boldsymbol{W}_j}[\sup_{\boldsymbol{\theta}_j} \|\boldsymbol{u}(\boldsymbol{W}_j, \boldsymbol{\theta}_j)\|] < \infty$$
$$\mathbb{E}_{\boldsymbol{W}_j}[\sup_{\boldsymbol{\theta}_j} \|\frac{\partial}{\partial \boldsymbol{\theta}_j}\boldsymbol{u}(\boldsymbol{W}_j, \boldsymbol{\theta}_j)\|] < \infty, \mathbb{E}_{\boldsymbol{W}_j}[\sup_{\boldsymbol{\theta}_j} \|\boldsymbol{u}(\boldsymbol{W}_j, \boldsymbol{\theta}_j)\boldsymbol{u}(\boldsymbol{W}_j, \boldsymbol{\theta}_j)^\top\|] < \infty$$
.

4. *For any* $\epsilon > 0$, $\inf_{\|\theta_j - \theta_j^\star\| \geq \epsilon} \|\mathbb{E}_{\boldsymbol{W}_j}[\boldsymbol{u}(\boldsymbol{W}_j, \boldsymbol{\theta}_j)]\| > 0 = \mathbb{E}_{\boldsymbol{W}_j}[\boldsymbol{u}(\boldsymbol{W}_j, \boldsymbol{\theta}_j^\star)]$

5. $\mathbb{E}_{\boldsymbol{W}_j}\left[\frac{\partial}{\partial \boldsymbol{\theta}}\boldsymbol{u}(\boldsymbol{W}_j, \boldsymbol{\theta}) \mid_{\boldsymbol{\theta}=\boldsymbol{\theta}_j^\star}\right]$ *is finite and positive definite.*

6. $\mathbb{E}_{\boldsymbol{W}_j}[\|u(\boldsymbol{W}_j, \boldsymbol{\theta}_j^\star)\|_3^3] < \infty$

*Proof.* We shall first establish the consistency of the estimator. By Assumption 3, we know there exists a positive integer $m$ such that $\boldsymbol{W}_{ij}$ and $\boldsymbol{W}_{(i+m)j}$ are independent for all $i \in \mathbb{N}$. Then for any $\boldsymbol{\theta}_j$, we know

$$\begin{aligned}
\operatorname{Var}\left(\frac{1}{N_{\text{int}}} \sum_{i \in D_{int}} \boldsymbol{u}(\boldsymbol{W}_{ij}, \boldsymbol{\theta}_j)\right) &= \frac{1}{n^2} \sum_{i \in D_{int}} \operatorname{Cov}\left(\boldsymbol{u}(\boldsymbol{W}_{ij}, \boldsymbol{\theta}_j), \sum_{k=-m+1}^{m-1} \boldsymbol{u}(\boldsymbol{W}_{(i+k)j}, \boldsymbol{\theta}_j)\right) \\
&\leq \frac{2m}{n^2} \sum_{i \in D_{int}} \operatorname{Cov}(\boldsymbol{u}(\boldsymbol{W}_{ij}, \boldsymbol{\theta}_j)) \\
&= \frac{2m}{n} \operatorname{Cov}(\boldsymbol{u}(\boldsymbol{W}_{ij}, \boldsymbol{\theta}_j)),
\end{aligned}$$

where the covariance is guaranteed to be finite by regularity condition 3. Therefore $\|\frac{1}{N_{\text{int}}} \sum_i \boldsymbol{u}(\boldsymbol{W}_{ij}, \boldsymbol{\theta}_j)\| = O_P(1/\sqrt{n})$ by Chebyshev's inequality. This establishes the pointwise convergence of $\boldsymbol{u}$. Uniform convergence then follows from regularity conditions 1, 2 and 3 as under these conditions the set of functions $\{\boldsymbol{u}(\cdot, \boldsymbol{\theta}) : \boldsymbol{\theta} \in \boldsymbol{\Theta}\}$ is Glivenko-Cantelli. By Theorem 5.9 of [5], the uniform convergence together with condition 4 ensures the consistency of $\widehat{\boldsymbol{\theta}}_j = (\widehat{\boldsymbol{b}}_j^\top, \widehat{\boldsymbol{\alpha}}_j^\top)^\top$. A similar argument establishes the consistency of $\widehat{\boldsymbol{\alpha}}_j'$.

Then we prove the asymptotic normality. In the previous analysis we showed that $\widehat{\boldsymbol{\theta}}_j$ converges in probability to $\boldsymbol{\theta}_j^\star$. Then combined with regularity condition 3, 5 , by Theorem 5.21 from [5] , we know

$$\begin{aligned}
\sqrt{n}(\widehat{\boldsymbol{\theta}} - \boldsymbol{\theta}^\star) &= -\mathbb{E}_{\boldsymbol{W}_j}\left[\frac{\partial}{\partial \boldsymbol{\theta}} \boldsymbol{u}(\boldsymbol{W}_j, \boldsymbol{\theta}) \mid_{\boldsymbol{\theta} = \boldsymbol{\theta}_j^\star}\right]^{-1} \frac{1}{\sqrt{n}} \sum_{i \in D_{int}} \boldsymbol{u}(\boldsymbol{W}_{ij}, \boldsymbol{\theta}^\star) + o_P(1) \\
&= -\begin{pmatrix} \boldsymbol{D}_1 & \boldsymbol{0} \\ \boldsymbol{0} & \boldsymbol{D}_2 \end{pmatrix}^{-1} \left(\frac{1}{\sqrt{n}} \sum_{i \in D_{int}} \boldsymbol{u}(\boldsymbol{W}_{ij}, \boldsymbol{\theta}^\star)\right) + o_P(1),
\end{aligned}$$

where the second equality follows from the definition of $\boldsymbol{u}, \boldsymbol{D}_1$ and $\boldsymbol{D}_2$. Similarly,

$$\begin{aligned}
\sqrt{n}(\widehat{\boldsymbol{\alpha}}' - \boldsymbol{\alpha}^\star) &= -\sqrt{\delta} \mathbb{E}_{\boldsymbol{W}_j}\left[\frac{\partial}{\partial \boldsymbol{\alpha}} \tilde{\boldsymbol{s}}(\boldsymbol{W}_j, \boldsymbol{\alpha}) \mid_{\boldsymbol{\alpha} = \boldsymbol{\alpha}_j^\star}\right]^{-1} \frac{1}{\sqrt{N}} \sum_{i \in D_{ext}} \tilde{\boldsymbol{s}}(\boldsymbol{W}_{ij}, \boldsymbol{\alpha}^\star) + o_P(1) \\
&= -\sqrt{\delta} \boldsymbol{D}_3^{-1} \left(\frac{1}{\sqrt{N}} \sum_{i \in D_{ext}} \tilde{\boldsymbol{s}}(\boldsymbol{W}_{ij}, \boldsymbol{\alpha}^\star)\right) + o_P(1)
\end{aligned}$$

By Assumption 3, $(\boldsymbol{u}(\boldsymbol{W}_{ij}, \boldsymbol{\theta}_j^\star))_{i \in D_{int}}$ and $(\tilde{\boldsymbol{s}}(\boldsymbol{W}_{ij}, \boldsymbol{\alpha}_j^\star))_{i \in D_{ext}}$ are m-dependent where $m$ is the maximum of family size in internal and external data. The asymptotic normality then follows from Theorem 3 of [6], under regularity condition 6. The proof is done by applying Slutsky's theorem. □

#### 4.3 Calculations under weak single-SNP effect

Assumption 5 also indicates that

$$\mathbb{E}\left[\boldsymbol{X}_{ij} \boldsymbol{X}_{i'j}^\top \mid (\boldsymbol{C}_i, \boldsymbol{C}_{i'})\right] \approx \mathbb{E}\left(\boldsymbol{X}_{ij} \boldsymbol{X}_{i'j}^\top\right), \quad \mathbb{E}\left[\boldsymbol{X}_{ij} \mid (\boldsymbol{C}_i, \boldsymbol{C}_{i'})\right] \approx \boldsymbol{0}.$$

We now approximate all the matrix elements $\boldsymbol{V}$ under Assumption 4 and Assumption 5. Denote $\mathrm{Var}(G_{ij}^2) = \sigma_{Gj}^2$ and $\mathrm{Cov}(\boldsymbol{X}_{ij}) = \boldsymbol{\Sigma}_{Gj}$. Then we have

$$\begin{aligned}
\boldsymbol{D}_1 = \mathbb{E}\left[\frac{\partial s_{ij}(\boldsymbol{b}_j^*)}{\partial \boldsymbol{b}_j}\right] &= -\mathbb{E}\left[g'\left(\boldsymbol{X}_{ij}^\top(\tau_j^*, \boldsymbol{\delta}_j^*) + \boldsymbol{C}_i^\top \boldsymbol{\gamma}_j^*\right)\begin{pmatrix}\boldsymbol{X}_{ij}\\ \boldsymbol{C}_i\end{pmatrix}\begin{pmatrix}\boldsymbol{X}_{ij}^\top & \boldsymbol{C}_i^\top\end{pmatrix}\right]\\
&= -\mathbb{E}\left[g'\left(\boldsymbol{C}_i^\top \boldsymbol{\gamma}_j^* + o(1)\right)\begin{pmatrix}\boldsymbol{X}_{ij}\\ \boldsymbol{C}_i\end{pmatrix}\begin{pmatrix}\boldsymbol{X}_{ij}^\top & \boldsymbol{C}_i^\top\end{pmatrix}\right]\\
&\approx -\mathbb{E}\left(\mathbb{E}\left[g'(\boldsymbol{C}_i^\top \boldsymbol{\gamma}_j^*)\begin{pmatrix}\boldsymbol{X}_{ij}\\ \boldsymbol{C}_i\end{pmatrix}\begin{pmatrix}\boldsymbol{X}_{ij}^\top & \boldsymbol{C}_i^\top\end{pmatrix} \mid \boldsymbol{C}_i\right]\right)\\
&\approx -\begin{pmatrix}\mathbb{E}\left[g'(\boldsymbol{C}_i^\top \boldsymbol{b}^*)\right]\boldsymbol{\Sigma}_{Gj} & \boldsymbol{0}\\ \boldsymbol{0} & \mathbb{E}\left[g'(\boldsymbol{C}_i^\top \boldsymbol{b}^*)\boldsymbol{C}_i\boldsymbol{C}_i^\top\right]\end{pmatrix}.
\end{aligned}$$

Similarly, we have

$$\begin{aligned}
\boldsymbol{D}_2 = \boldsymbol{D}_3 &\approx -\begin{pmatrix}\mathbb{E}\left[g'(\boldsymbol{C}_i^\top \boldsymbol{\alpha}_{2j}^*)\right]\sigma_{Gj}^2 & \boldsymbol{0}\\ \boldsymbol{0} & \mathbb{E}\left[g'(\boldsymbol{C}_i^\top \boldsymbol{\alpha}_{2j}^*)\boldsymbol{C}_i\boldsymbol{C}_i^\top\right]\end{pmatrix}\\
&\approx -\begin{pmatrix}\mathbb{E}\left[g'(\boldsymbol{C}_i^\top \boldsymbol{b}^*)\right]\sigma_{Gj}^2 & \boldsymbol{0}\\ \boldsymbol{0} & \mathbb{E}\left[g'(\boldsymbol{C}_i^\top \boldsymbol{b}^*)\boldsymbol{C}_i\boldsymbol{C}_i^\top\right]\end{pmatrix}.
\end{aligned}$$

Additionally, for any two individuals $i$ and $i'$ (including $i = i'$), we have the approximation:

$$\begin{aligned}
&\mathrm{Cov}\left[s_{ij}(\boldsymbol{b}_j^*), s_{i'j}(\boldsymbol{b}_j^*)\right] = \mathbb{E}\left[s_{ij}(\boldsymbol{b}_j^*)s_{i'j}(\boldsymbol{b}_j^*)^\top\right]\\
&= \mathbb{E}\left[(Y_i - \mu_{ij})(Y_{i'} - \mu_{i'j})\begin{pmatrix}\boldsymbol{X}_{ij}\\ \boldsymbol{C}_i\end{pmatrix}\begin{pmatrix}\boldsymbol{X}_{i'j}^\top & \boldsymbol{C}_{i'}^\top\end{pmatrix}\right]\\
&\approx \mathbb{E}\left[(Y_i - g(\boldsymbol{C}_i^\top \boldsymbol{b}_{3j}^*))(Y_{i'} - g(\boldsymbol{C}_{i'}^\top \boldsymbol{b}_{3j}^*))\begin{pmatrix}\boldsymbol{X}_{ij}\\ \boldsymbol{C}_i\end{pmatrix}\begin{pmatrix}\boldsymbol{X}_{i'j}^\top & \boldsymbol{C}_{i'}^\top\end{pmatrix}\right]\\
&\approx -\begin{pmatrix}\mathbb{E}\left[(Y_i - g(\boldsymbol{C}_i^\top \boldsymbol{b}^*))(Y_{i'} - g(\boldsymbol{C}_{i'}^\top \boldsymbol{b}^*))\right]\mathbb{E}\left(\boldsymbol{X}_{ij}\boldsymbol{X}_{i'j}^\top\right) & \boldsymbol{0}\\ \boldsymbol{0} & \mathbb{E}\left[(Y_i - g(\boldsymbol{C}_i^\top \boldsymbol{b}^*))(Y_{i'} - g(\boldsymbol{C}_{i'}^\top \boldsymbol{b}^*))\boldsymbol{C}_i\boldsymbol{C}_{i'}^\top\right]\end{pmatrix}
\end{aligned}$$

And similarly,

$$\begin{aligned}
&\mathrm{Cov}\left[s_{ij}(\boldsymbol{b}_j^*), \tilde{s}_{i'j}(\boldsymbol{a}_j^*)\right] = \mathbb{E}\left[s_{ij}(\boldsymbol{b}_j^*)\tilde{s}_{i'j}(\boldsymbol{a}_j^*)^\top\right]\\
&\approx -\begin{pmatrix}\mathbb{E}\left[(Y_i - g(\boldsymbol{C}_i^\top \boldsymbol{b}^*))(Y_{i'} - g(\boldsymbol{C}_{i'}^\top \boldsymbol{b}^*))\right]\mathbb{E}\left(\boldsymbol{X}_{ij}G_{i'j}\right) & \boldsymbol{0}\\ \boldsymbol{0} & \mathbb{E}\left[(Y_i - g(\boldsymbol{C}_i^\top \boldsymbol{b}^*))(Y_{i'} - g(\boldsymbol{C}_{i'}^\top \boldsymbol{b}^*))\boldsymbol{C}_i\boldsymbol{C}_{i'}^\top\right]\end{pmatrix}
\end{aligned}$$

and

$$\begin{aligned}
&\mathrm{Cov}\left[\tilde{s}_{ij}(\boldsymbol{a}_j^*), \tilde{s}_{i'j}(\boldsymbol{a}_j^*)\right] = \mathbb{E}\left[\tilde{s}_{ij}(\boldsymbol{a}_j^*)\tilde{s}_{i'j}(\boldsymbol{a}_j^*)^\top\right]\\
&\approx -\begin{pmatrix}\mathbb{E}\left[(Y_i - g(\boldsymbol{C}_i^\top \boldsymbol{b}^*))(Y_{i'} - g(\boldsymbol{C}_{i'}^\top \boldsymbol{b}^*))\right]\mathbb{E}\left(G_{ij}G_{i'j}\right) & \boldsymbol{0}\\ \boldsymbol{0} & \mathbb{E}\left[(Y_i - g(\boldsymbol{C}_i^\top \boldsymbol{b}^*))(Y_{i'} - g(\boldsymbol{C}_{i'}^\top \boldsymbol{b}^*))\boldsymbol{C}_i\boldsymbol{C}_{i'}^\top\right]\end{pmatrix}
\end{aligned}$$

#### 4.4 Variance reduction when $\mathbf{F}_{ij} = (G_{ij}^f, G_{ij}^m)$.

Consider the case when the family genotype $\boldsymbol{F}_{ij} = (G_{ij}^f, G_{ij}^m)$ and the model is

$$g(\mathbb{E}[Y_i \mid G_{ij}, \boldsymbol{F}_{ij}, \boldsymbol{C}_i]) = \tau_j G_{ij} + \delta_{1j}G_{ij}^f + \delta_{2j}G_{ij}^m + \boldsymbol{\gamma}_j^\top \boldsymbol{C}_i$$

We know $\mathrm{Cov}(G_{ij}, G_{ij}^f + G_{ij}^m) = \mathrm{Cov}(G_{ij}, T_{ij} + NT_{ij}) = \sigma_{G_j}^2(1 + r_j)$. By symmetry,

$$\mathrm{Cov}(G_{ij}, G_{ij}^f) = \mathrm{Cov}(G_{ij}, G_{ij}^m) = \sigma_{G_j}^2(1 + r_j)/2.$$

Similarly, we know $\text{Cov}(G_{ij}^f + G_{ij}^m, G_{ij}^f + G_{ij}^m) = \text{Var}(T_{ij} + NT_{ij}) = 2\sigma_{G_j}^2(1+r_j)$. Therefore

$$\text{Cov}(G_{ij}^f, G_{ij}^m) = \frac{\text{Cov}(G_{ij}^f + G_{ij}^m, G_{ij}^f + G_{ij}^m) - \text{Var}(G_{ij}^f) - \text{Var}(G_{ij}^m)}{2} = \sigma_{G_j}^2 r_j$$

Hence we have

$$\Sigma_{G_j} = \sigma_{Gj}^2 \begin{pmatrix} 1 & (1+r_j)/2 & (1+r_j)/2 \\ (1+r_j)/2 & 1 & r_j \\ (1+r_j)/2 & r_j & 1 \end{pmatrix}, \quad \text{Cov}(\boldsymbol{X}_{ij}, G_{ij}) = \sigma_{Gj}^2 \begin{pmatrix} 1 \\ (1+r_j)/2 \\ (1+r_j)/2 \end{pmatrix}$$

It follows that

$$\Sigma_{G_j}^{-1} = \sigma_{G_j}^{-2} \begin{pmatrix} \frac{2}{1-r_j} & -\frac{1}{1-r_j} & -\frac{1}{1-r_j} \\ -\frac{1}{1-r_j} & \frac{1}{2}\frac{r_j+3}{1-r_j^2} & \frac{1}{2}\frac{1}{1+r_j} \\ -\frac{1}{1-r_j} & \frac{1}{2}\frac{1}{1+r_j} & \frac{1}{2}\frac{r_j+3}{1-r_j^2} \end{pmatrix}$$

Then

$$\text{Cov}\left(\begin{pmatrix} \widehat{\tau}_j \\ \widehat{\delta}_{1j} \\ \widehat{\delta}_{2j} \\ \widehat{\alpha}_j^{\text{int}} \\ \widehat{\alpha}_1^{\text{ext}} \end{pmatrix}, \begin{pmatrix} \widehat{\tau}_j \\ \widehat{\delta}_{1j} \\ \widehat{\delta}_{2j} \\ \widehat{\alpha}_j^{\text{int}} \\ \widehat{\alpha}_j^{\text{ext}} \end{pmatrix}\right)$$

$$\approx \frac{\sigma_{Y|C}^2}{N_{\text{int}}\mathbb{E}\left[g'(\boldsymbol{C}_i^T\boldsymbol{b}^*)\right]^2 \sigma_{Gj}^2} \begin{pmatrix} \frac{2}{1-r_j} & -\frac{1}{1-r_j} & -\frac{1}{1-r_j} & 1 & \frac{N_{\text{share}}}{N_{\text{ext}}} \\ -\frac{1}{1-r_j} & \frac{1}{2}\frac{r_j+3}{1-r_j^2} & \frac{1}{2}\frac{1}{1+r_j} & 0 & 0 \\ -\frac{1}{1-r_j} & \frac{1}{2}\frac{1}{1+r_j} & \frac{1}{2}\frac{r_j+3}{1-r_j^2} & 0 & 0 \\ 1 & 0 & 0 & 1 & \frac{N_{\text{share}}}{N_{\text{ext}}} \\ \frac{N_{\text{share}}}{N_{\text{ext}}} & 0 & 0 & \frac{N_{\text{share}}}{N_{\text{ext}}} & \frac{N_{\text{int}}}{N_{\text{ext}}} \end{pmatrix}$$

This implies that

$$\text{Var}\left(\begin{pmatrix} \widehat{\tau}_j \\ \widehat{\alpha}_{1j} - \widehat{\alpha}'_{1j} \end{pmatrix}\right) = \text{Var}\left(\begin{pmatrix} \widehat{b}_{1j} \\ \widehat{\alpha}_{1j} - \widehat{\alpha}'_{1j} \end{pmatrix}\right) = \frac{\sigma_{Y|C}^2}{N_{\text{int}}\mathbb{E}\left[g'(\boldsymbol{C}_i^T\boldsymbol{b}^*)\right]^2 \sigma_{Gj}^2} \begin{pmatrix} \frac{2}{1-r_j} & 1 - \frac{N_{\text{share}}}{N_{\text{ext}}} \\ 1 - \frac{N_{\text{share}}}{N_{\text{ext}}} & 1 + \frac{N_{\text{int}}}{N_{\text{ext}}} - \frac{2N_{\text{share}}}{N_{\text{ext}}} \end{pmatrix}$$

This matches the result obtained when $\boldsymbol{F}_{ij} = G_{ij}^{pa}$, and the remainder of the variance reduction proof proceeds identically to that case.

#### 5 Supplemental Figures

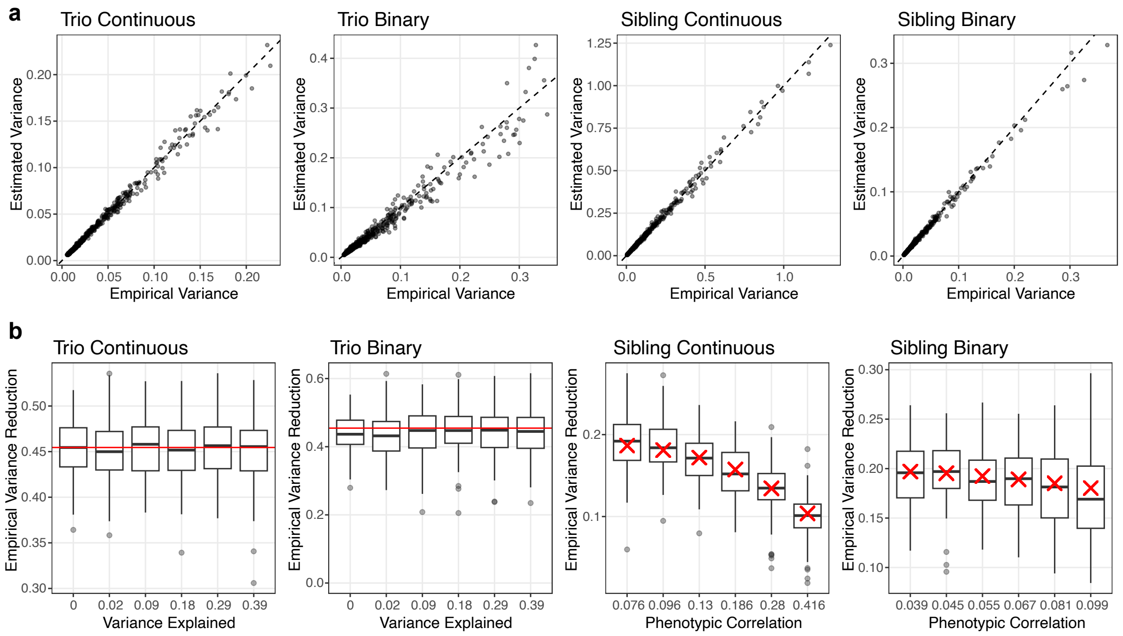


Figure S1: Simulation results using only summary statistics. a) Comparison of the estimated variance of the calibrated estimator using summary statistics with its empirical variance across 500 simulation replicates. Each black dot represents one SNP, and the diagonal line denotes $y = x$. b) Empirical variance reduction of the calibrated estimator using summary statistics relative to the uncalibrated estimator for 100 SNPs under linear (continuous-trait) and logistic (binary-trait) regression in trio and sibling designs. Each black dot represents one SNP; boxplots summarize the distribution (median, interquartile range, and whiskers showing the central 1.5 IQR range) across SNPs. Red lines or crosses denote the corresponding theoretical variance reduction.

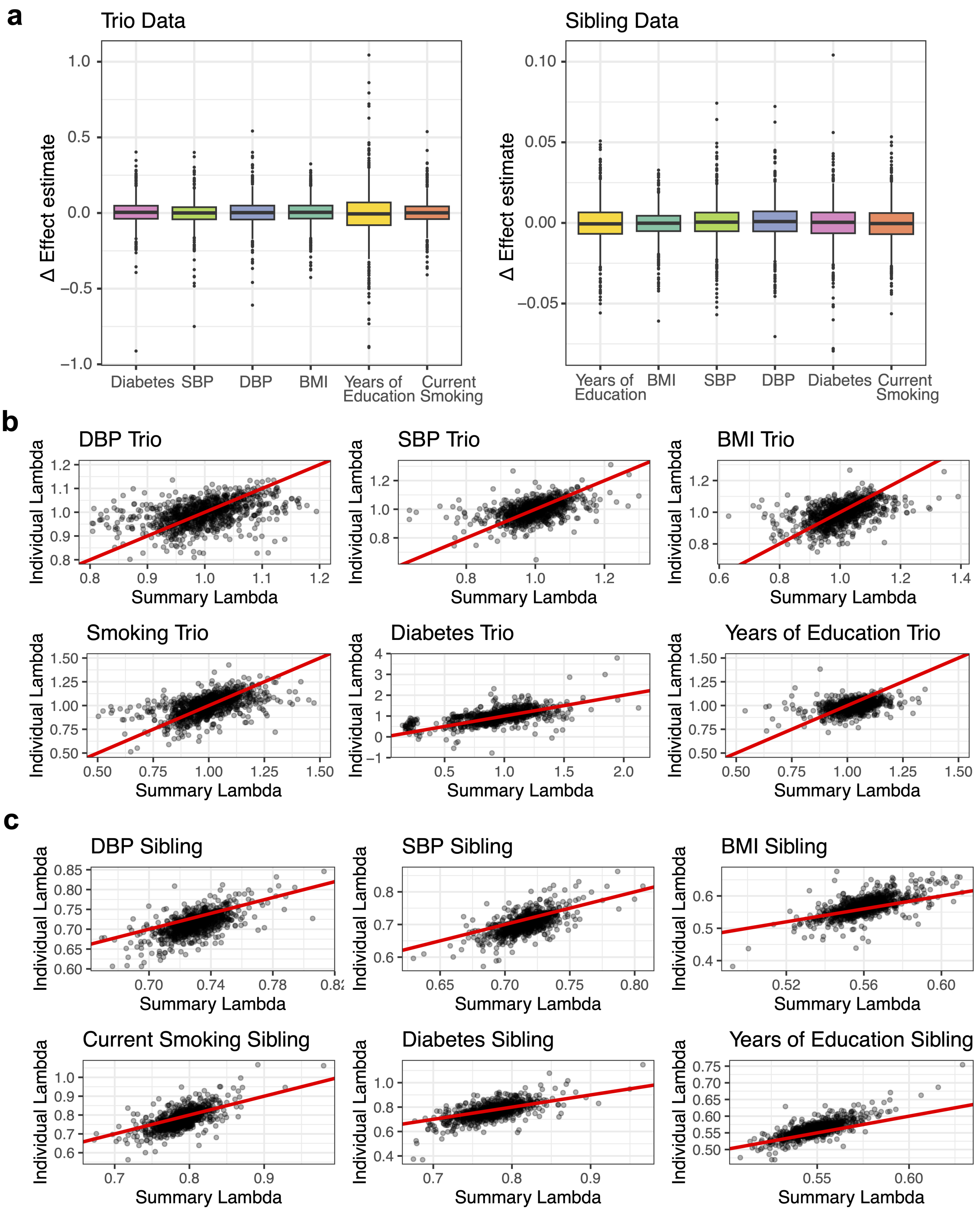


Figure S2: Additional results for calibration applied to UK Biobank traits. a) Differences of effect estimates between the calibrated and uncalibrated estimators when used on trio and sibling data. The boxplots are based on 1,000 randomly sampled SNPs and summarize the distribution of the difference of effect estimates under the trio and sibling designs (median, interquartile range, and whiskers showing the central 1.5 IQR range). b) Scatter plots of $\widehat{\lambda}_j^*$ computed from individual-level data and summary-level data of 1000 randomly sampled SNPs. Each dot is a SNP. The red lines represent the diagonal line $y = x$.

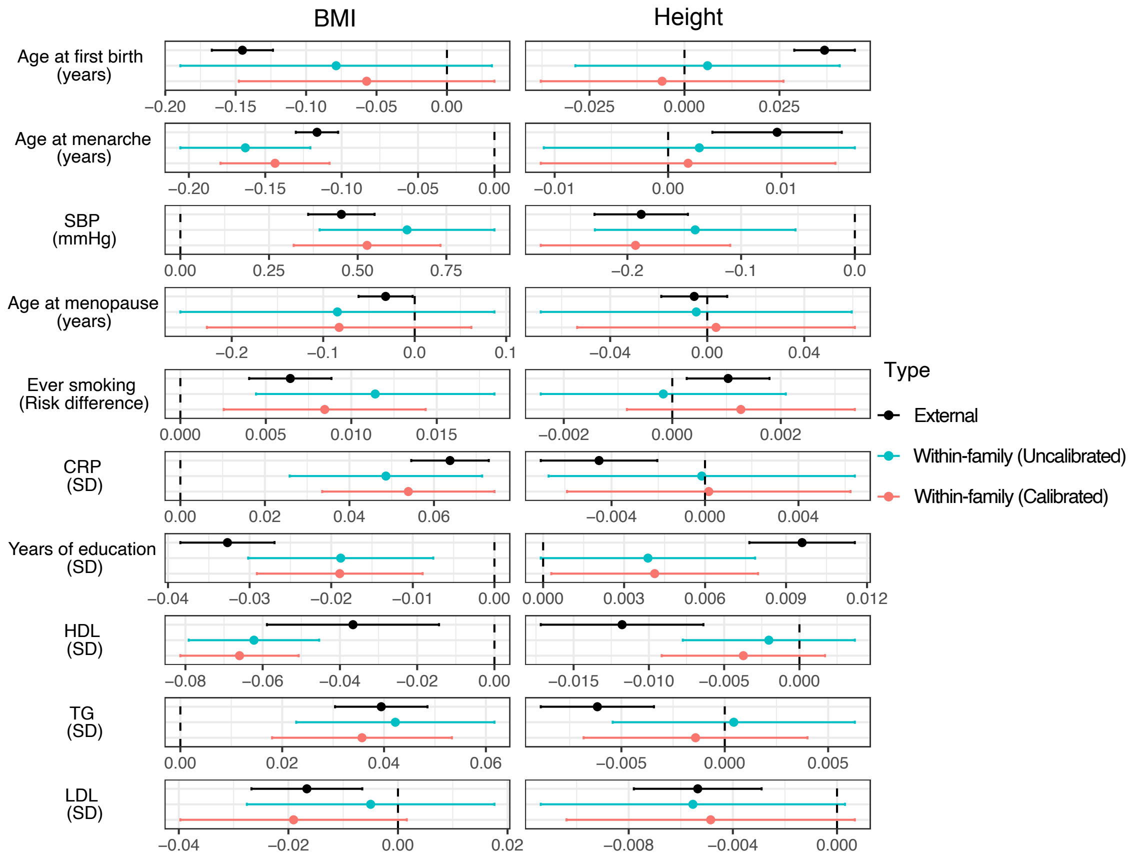


Figure S3: Calibration applied to published within-sibship GWAS summary statistics. MR estimates with IVW for BMI and height using three sets of summary statistics from published within-sibship GWAS: population-based (external GWAS), within-family uncalibrated, and within-family calibrated. Point estimates are shown with 95% confidence intervals, computed using IVW.
